# Status and Opportunities of Machine Learning Applications in Obstructive Sleep Apnea: A Narrative Review

**DOI:** 10.1101/2025.02.27.25322950

**Authors:** Matheus Lima Diniz Araujo, Trevor Winger, Samer Ghosn, Carl Saab, Jaideep Srivastava, Louis Kazaglis, Piyush Mathur, Reena Mehra

**Affiliations:** Cleveland Clinic Foundation, Cleveland, OH, USA; University of Minnesota, Minneapolis, MN, USA; Epic Systems Corporation, Verona, WI, USA; USA University of Washington, Seattle, WA, USA

**Keywords:** Sleep Apnea, Machine Learning

## Abstract

**Background:** Obstructive sleep apnea (OSA) is a prevalent and potentially severe sleep disorder characterized by repeated interruptions in breathing during sleep. Machine learning models have been increasingly applied in various aspects of OSA research, including diagnosis, treatment optimization, and developing biomarkers for endotypes and disease mechanisms.

**Objective:** This narrative review evaluates the application of machine learning in OSA research, focusing on model performance, dataset characteristics, demographic representation, and validation strategies. We aim to identify trends and gaps to guide future research and improve clinical decision-making that leverages machine learning.

**Methods:** This narrative review examines data extracted from 254 scientific publications published in the PubMed database between January 2018 and March 2023. Studies were categorized by machine learning applications, models, tasks, validation metrics, data sources, and demographics.

**Results:** Our analysis revealed that most machine learning applications focused on OSA classification and diagnosis, utilizing various data sources such as polysomnography, electrocardiogram data, and wearable devices. We also found that study cohorts were predominantly overweight males, with an underrepresentation of women, younger obese adults, individuals over 60 years old, and diverse racial groups. Many studies had small sample sizes and limited use of robust model validation. Conclusion: Our findings highlight the need for more inclusive research approaches, starting with adequate data collection in terms of sample size and bias mitigation for better generalizability of machine learning models in OSA research. Addressing these demographic gaps and methodological opportunities is critical for ensuring more robust and equitable applications of artificial intelligence in healthcare.

## Introduction

Obstructive sleep apnea (OSA) is a highly prevalent condition worldwide, affecting an estimated 46% to 62% of the adult global population [1], with approximately 1 billion adults having mild to severe conditions and 425 million adults aged 30-69 years having moderate to severe OSA [2]. Prevalence varies by region and demographic characteristics. For example, in China, 8.8% of the adult population is estimated to have moderate to severe sleep apnea, while in Brazil, the estimate is 26% of the population[2]. In a large population-based study conducted in Switzerland, 49.7% of the men and 23.4% of the women had moderate to high sleep-disordered breathing [3]. OSA also affects a significant portion of the US population, with approximately 38% of the US population being diagnosed with mild severity and up to 17% diagnosed with moderate severity [3]. These figures highlight the widespread burden of OSA and underscore the need for scalable, efficient diagnostic and treatment strategies.

OSA is characterized by frequent, intermittent cessation (apnea) or reduction (hypopnea) of airflow during sleep, and it can severely affect an individual’s health if left untreated, with serious negative ramifications for global health if unrecognized and untreated [4]. The gold standard for sleep apnea diagnosis is polysomnography (PSG), which records a large amount of patient data while sleeping, including brain waves, blood oxygen levels, heart rate, breathing, and eye and leg movements [5]. Home Sleep Apnea Tests (HSATs) have emerged as practical diagnostic tools since patients can perform their sleep study at home. However, the HSAT approach is standardly less comprehensive than the in-laboratory PSG. A key diagnostic metric is the Apnea-Hypopnea Index (AHI), calculated based on the number of apneas and hypopnea events per hour of sleep derived from the sleep study. Although AHI tends to oversimplify the complex nature of individual OSA cases, clinicians widely use this metric, which traditionally considers 5, 15, and 30 events/hour thresholds to discriminate normal, mild, moderate, and severe cases, respectively [6], with diagnosis integrating the clinical presentation of the patient’s symptoms and comorbidities [7].

In sleep medicine, the study, diagnosis, prognosis, and treatment of OSA pose numerous challenges, from treatment adherence to diagnostic metrics. At the same time, OSA research provides uniquely rich data regarding demographic representation and high-density physiological signals that, when leveraged by innovative technologies such as applied machine learning and wearable devices, can address current OSA challenges and contribute to translational research across different medical domains [8].

Machine learning can extract complex patterns from data to perform classification and regression tasks, which is critical to reveal hidden knowledge in large volumes of data. For example, when seeking a new, low-cost, and feasible OSA diagnosis tool, Wang et al. (2022), used a deep learning algorithm based on sleep sounds for apnea detection. In another innovative task, Araujo et al. (2019) used a large cohort of thousands of patients receiving continuous positive airway pressure (CPAP) therapy for OSA management to train machine-learning models to predict when interventions are needed due to low adherence [9, 10].

Several reviews have targeted machine learning applications in OSA. In one study, Mendonca et al. (2019) focused on analyzing algorithms for sleep apnea diagnoses based on pulse oximetry, electrocardiogram (ECG), and sound analyses [11]. In another study, Ramachandran et al. (2021) searched the literature for different types of sensors used for data collection and feature engineering techniques for sleep apnea detection [12]. Similarly, Ferreira-Santos et al. (2022) conducted a comprehensive evaluation of clinical prediction algorithms for OSA diagnosis. They emphasized the lack of external validation and the high risk of bias and applicability in most studies, despite the frequent use of simple predictors like BMI, age, and sex [13]. Despite these reviews, no review to our knowledge has made a broader investigation into different applications of machine learning beyond OSA diagnosis, and a detailed analysis of the different data type modalities, demographic discrepancies, machine learning approaches, and evaluation metrics.

Our objectives for this review are to re-evaluate current data modeling methodologies, understand potential existing biases, and chart the way forward using traditional machine learning in OSA research.

## Methods

We conducted our review by initially identifying search terms that align with our purpose and research questions, employing an iterative approach to select studies and manually extract data. We then performed a numerical summary of key data elements, followed by a report on the results and a discussion of the findings with specific attention to their implications for clinical practice, research, and future investigations into machine learning applications in OSA.

Our literature review used PubMed’s (National Library of Medicine, Bethesda, MD) advanced search to identify studies released from January 2018 to March 2023. PubMed is a comprehensive and authoritative database widely used in biomedical research, ensuring access to high-quality, peer-reviewed articles relevant to obstructive sleep apnea and machine learning [14].

Our research criteria were to include original, peer-reviewed studies published between January 2018 and March 2023 that explicitly applied machine learning techniques to OSA. Articles had to be written in English, involving human subjects, and focusing on OSA, excluding studies centered on central or mixed sleep apnea. We also excluded animal studies and non-original research formats such as reviews, surveys, editorials, and commentaries.

We searched for machine learning-related terms associated with OSA for the search criteria, resulting in following search query: *(((‘Machine-learning’) OR (‘Artificial-intelligence’) OR (‘Neural-networks’) OR (‘Deep-learning’) OR (‘Reinforcement-learning’) OR (‘Predictive-modeling’) OR (‘Statistical-learning’) OR (‘Computer-vision’) OR (‘Natural-language-processing’) OR (‘Decision-trees’) OR (‘Random-forests’) OR (‘Support-vector-machines’) OR (‘Clustering’) OR (‘Convolutional-neural-networks’) OR (‘Recurrent-neural-networks’) OR (‘Generative-adversarial-networks’) OR (‘Transfer-learning’) OR (‘Ensemble-learning’) OR (‘Unsupervised-learning’) OR (‘Supervised-learning’) OR (‘Semi-supervised-learning’) OR (‘Active-learning’) OR (‘Bayesian-networks’) OR (‘Decision-tree’) OR (‘Random-forest’) OR (‘Support-vector-machines’) OR (‘SVM’) OR (‘Naive-Bayes’) OR (‘K-nearest-neighbors’) OR (‘KNN’) OR (‘Principal-component-analysis’) OR (‘PCA’) OR (‘Recurrent-neural-networks’) OR (‘RNN’) OR (‘Convolutional-neural-networks’) OR (‘CNN’) OR (‘Generative-adversarial-networks’) OR (‘GANs’) OR (‘Autoencoders’) OR (‘Hidden-Markov-models’) OR (‘HMMs’) OR (‘Long-short-term-memory’) OR (‘LSTM’)) AND ((‘obstructive-sleep-apnea’)*.

Our query returned 586 references using the search criteria. These references underwent three filtering processes, as shown in Figure 1. First, we removed 180 duplicate references. Following our inclusion and exclusion criteria, we excluded 124 studies that were not relevant based on title and abstract analysis. Finally, we removed 27 studies after analyzing the complete text because they were not original publications, did not use machine learning in their methodology, were non-English publications, or were related to central or mixed sleep apnea instead of OSA. The final dataset for data extraction included 254 studies. For the screening and data extraction process, we used Covidence^1^ software (Veritas Health Innovation, Brooklyn, NY). The full reference of each included study is available in the supplemental materials.

**Figure 1:**
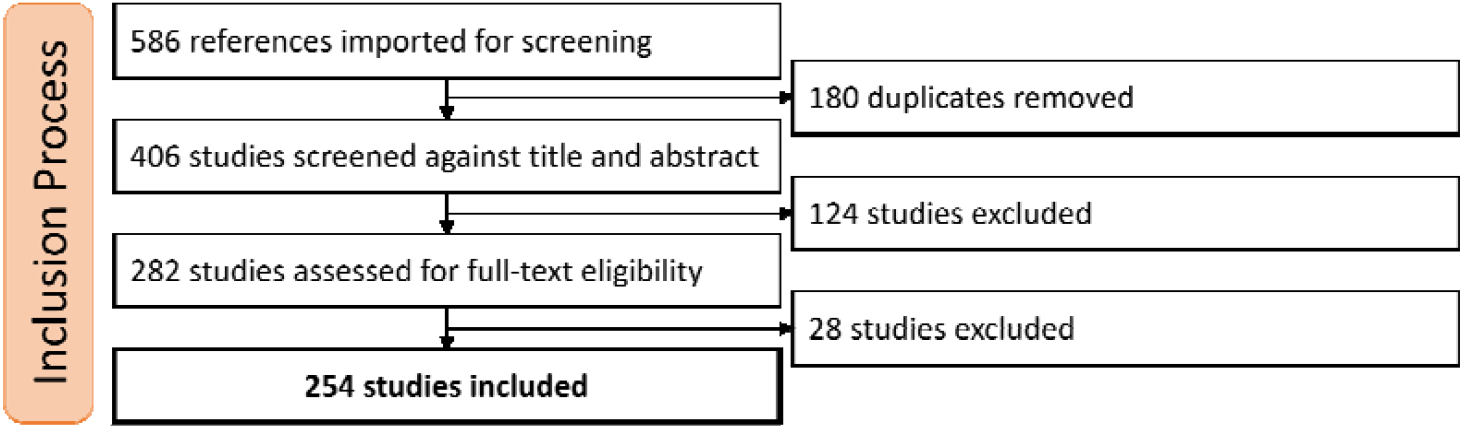
Inclusion process diagram. Diagram showing the survey including process with 3 major filtering steps.

We performed manual data extraction from the full text of each study with subsequent second-person revision. While screening the text, our group extracted the main application of machine learning from each article, categorizing them into 18 major subjects. We extracted the following machine learning-related variables: the type of model used (e.g., logistic regression, random forest), task type (e.g., classification, regression), and traditional validation metrics (e.g., accuracy, precision, recall, area under the ROC curve). We also extracted information about the data source (e.g., polysomnogram, questionnaires, electronic health records) and the data type (e.g., tabular data, raw audio). Moreover, demographic sample information was also extracted (e.g., age, sex, race, BMI). We removed studies that did not have the reported information for each analysis. This data extraction and categorization process enabled our quantitative thematic analysis.

## Results

### Analysis of Studies Subject Categories

Table 1 presents the distribution of 18 machine-learning application topics in OSA research. The primary focus of the studies, representing 30% (77 studies), was on using machine learning to classify sleep apnea diagnosis. Other significant categories include the automatic identification of hypopnea/apnea events at the epoch level (11%) and the detection and profiling of snoring (10%). Less common applications involve the severity stratification of sleep apnea using the Apnea-Hypopnea Index (AHI) (8%), the prediction of mortality and cardiovascular risks (7%), and the automatic classification of sleep stages (7%), reflecting the diverse potential of machine learning in enhancing diagnostic and predictive accuracy in sleep medicine. The table shows the broad spectrum of research, including categories focusing on novel technologies (e.g., wearable devices) for diagnosis, machine learning in genetic screening, and treatment-related applications (e.g., CPAP adherence) [15-17].

**Table 1:** Absolute and relative popularity of studies categories. One study can have more than one study category.

| Machine Learning Applications | Number of studies | Proportion (N=254) |
| --- | --- | --- |
| Sleep apnea diagnosis classification | 74 | 29% |
| Identification of hypopnea/apnea at epoch level | 29 | 11% |
| Snoring detection and profiling | 25 | 10% |
| Severity stratification (AHI) | 20 | 8% |
| Predict mortality risk, cardiovascular risk, and other conditions related to Sleep Apnea | 19 | 7% |
| Automatic classification of sleep stage in sleep apnea | 19 | 7% |
| Detecting or screening other conditions related to Sleep apnea (e.g adenoid hypertrophy) | 16 | 6% |
| Wearable or nearable devices | 15 | 6% |
| Screening and profiling of protein/DNA/genetic/blood analysis for sleep apnea | 14 | 5% |
| Clustering analysis for phenotyping those with sleep apnea | 13 | 5% |
| OSA questionnaires analysis | 9 | 4% |
| CPAP adherence and usage | 8 | 3% |
| Electronic Health Records (EHR) analysis | 7 | 3% |
| Sleep posture classification | 5 | 2% |
| Predicting treatment outcomes | 3 | 1% |
| Predicting candidate for surgery | 3 | 1% |
| Predicting score results for drug-induced sleep endoscopy | 3 | 1% |
| Obstructive Sleep Apnea vs Central Sleep Apnea | 3 | 1% |

### Demographic analysis of age, sex, body mass index, and race

We recorded several key demographic characteristics that provided insight into the participant profiles across various studies, including age, sex, BMI, and race.

The age distribution of the study population reveals a distinct bimodal pattern, as shown in Figure 2a. This pattern highlights a primary focus on pediatric research and individuals who are 40-60 years old.

**Figure 2:**
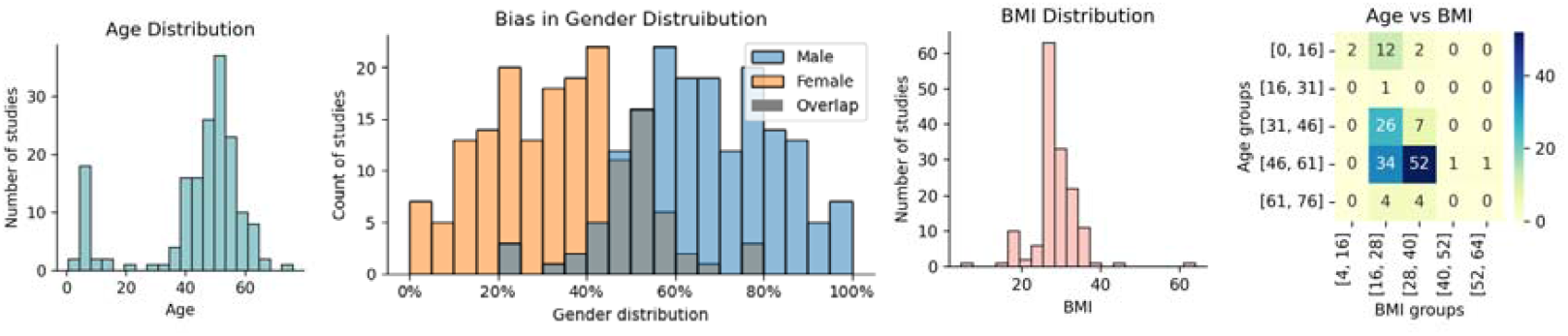
Mean demographic data distribution. (a) Mean age distribution, (b) gender distribution across publications highlighting gender bias, (c) mean BMI distribution and (d) heatmap showing the number of studies by BMI and age groups.

The participant’s BMI across various studies was 28.2±6.18kg/m^2^, indicating a prevalent trend toward the overweight but not severely obese population. The presence of extreme values, as seen in Figure 2c, suggests variability in the health profiles of the participants, which may include the pediatric population with relatively low BMI. The heatmap of both mean age and BMI groups in Figure 2d indicates potential gaps in research, such as a low representation of studies with younger obese adults and low representation of older populations (i.e., populations with a higher prevalence of OSA).

Sex-specific distributions varied significantly across studies, as shown by the sex distribution histograms in Figure 2b. On average, males constituted 67±16% of the samples, in contrast with females, who constituted an average of 33±16%, which is consistent with the higher prevalence of OSA in men versus women.

The studies span multi-national geographical locations, including countries across Asia, Europe, and America. Race or ethnicity is mentioned in 142 studies (55%), but only 20 (8%) explicitly provided a multiracial or multiethnic sample participation description.

### Analysis of Data Source and Types

We distinguished between the sources of data and their respective types. The source indicates the high-level origin of data, and they included *-omics analysis, polysomnography (PSG), medical devices, electronic health records (EHR), questionnaires, wearables and other devices, positive airway pressure (PAP)-devices, blood analysis, and microphones. Each source may encompass multiple data types. For instance, in a polysomnogram, data collected include electrocardiogram (ECG), oxygen saturation (SpO2), and electroencephalogram (EEG) types. The distribution of each data source and type across the studies is detailed in Figure 3.

**Figure 3:**
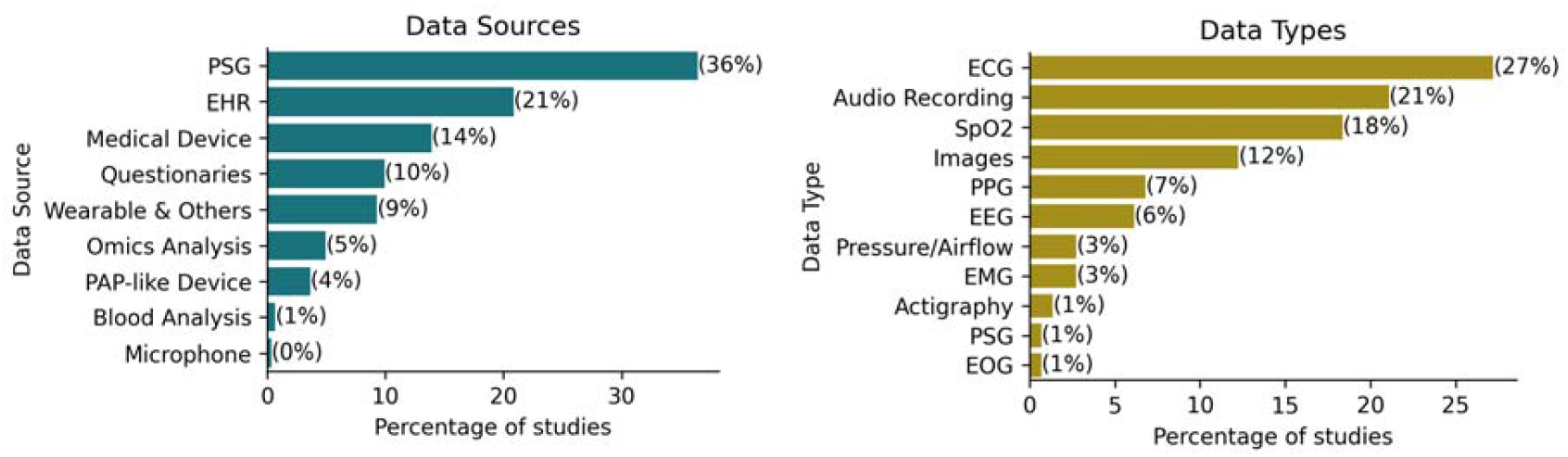
Popularity of Data Types (left) and Data Sources (right).

We found that 83 studies (32%) utilized multiple data sources, underscoring the multimodal nature of data integration in OSA research. The most used data type was ECG, which appeared in 27% of the studies, which was mostly derived from PSG (36%). Following PSG, EHR was utilized in 21% of the studies. Interestingly, despite representing the primary treatment for OSA, data from Positive Airway Pressure (PAP) devices were available in only 4% of the studies.

### Choice of machine learning algorithms over time

We assessed the popularity of machine learning models over the years( Figure 4 -Top). Deep Learning models are the most popular, followed by support vector machines, which have a negative trend, and Ensemble methods like XGBoost and Random Forest models, which have an increasing trend. Traditional linear models have remained consistent in popularity over the years.

**Figure 4:**
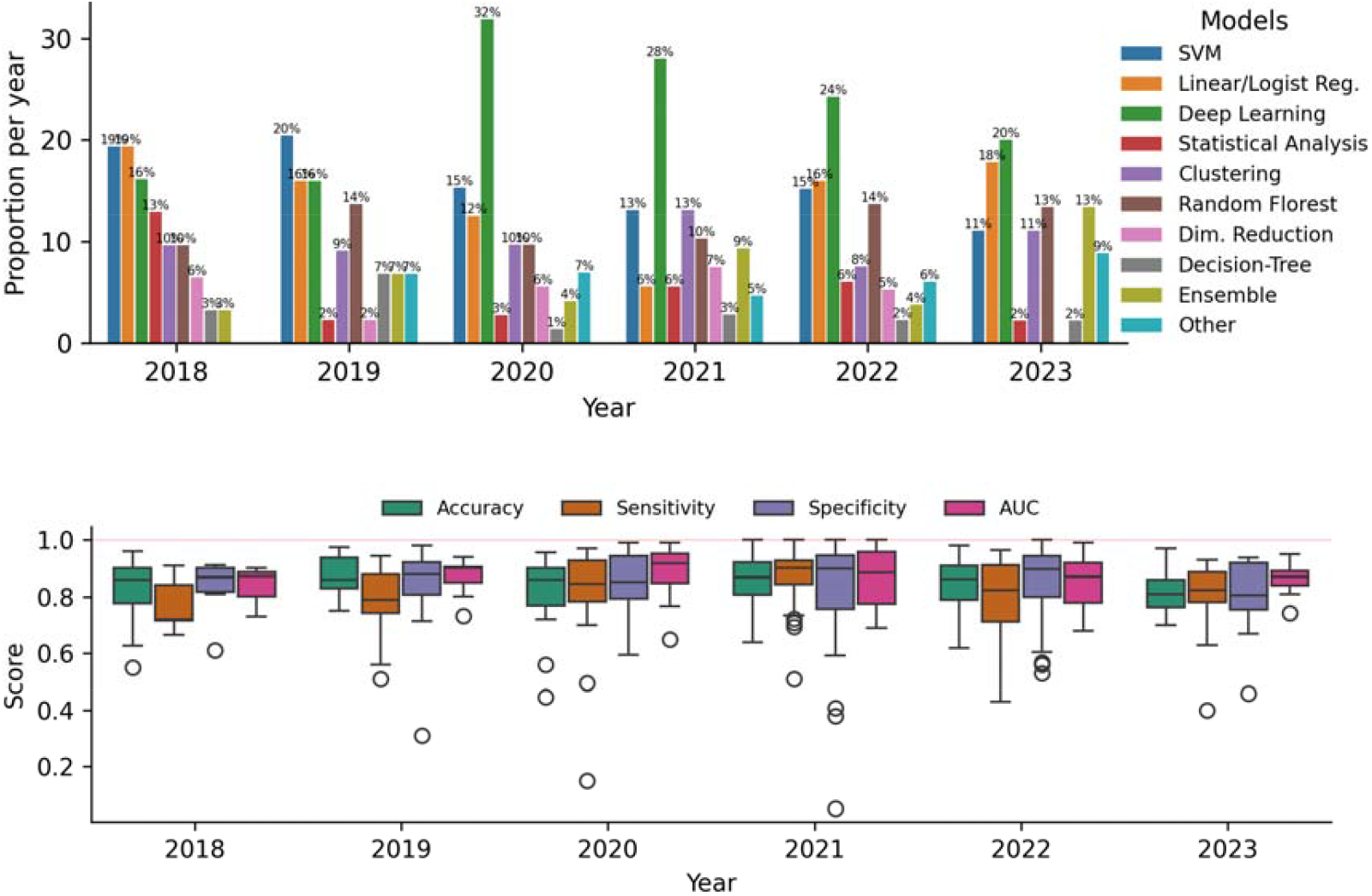
(Top) Bar plot showing yearly proportions of machine learning model usage from 2018 to 2023, highlighting trends in the popularity of different model types over time. (Bottom) Boxplots of aggregate performance metrics (Accuracy, Sensitivity, Specificity, AUC) from 2018 to 2023, illustrating trends in model evaluation scores over time.

### Analysis of Classification Evaluation Metrics

We evaluated the types of machine learning tasks addressed in each study, focusing on classification, clustering, and regression. Classification was the most common task, appearing in 195 studies. Given this prevalence, we conducted a detailed analysis of classification-related performance metrics. The most frequently reported metrics were accuracy (163 studies), sensitivity (127 studies), specificity (123 studies), and area under the receiver operating characteristic curve (AUC, 106 studies). Figure 4 presents the distribution of these metric scores over the years, which typically ranged from 0.78 to 0.92, with an average of 0.83. Papers published in 2018 showed comparatively lower scores than those in subsequent years. Additionally, some papers from 2020, 2021, and 2022 reported perfect scores of 100%.

### Sample Size, Test Size, and Validation Strategy

Our analysis indicates that a wide range of sample sizes are used as datasets for machine learning applied to OSA. In Figure 5 (a), we show the logarithmic distribution of the sample size, where we can highlight that most studies (77%) have sample sizes up to 1000 samples. Only 6 studies utilized a sample size of more than 10000 samples, and only one study used more than 1 million samples, which used a large sample to determine the most critical predictor of time to death between central and obstructive sleep apnea groups [18]. Additionally, Figure 5(b) shows the proportion of data allocated for testing the machine learning models. We found that the majority of studies reserved up to 30% of their data for testing. The most common test size was 20%, used in 28 studies, followed by 30% in 26 studies and 50% in 18 studies.

**Figure 5:**
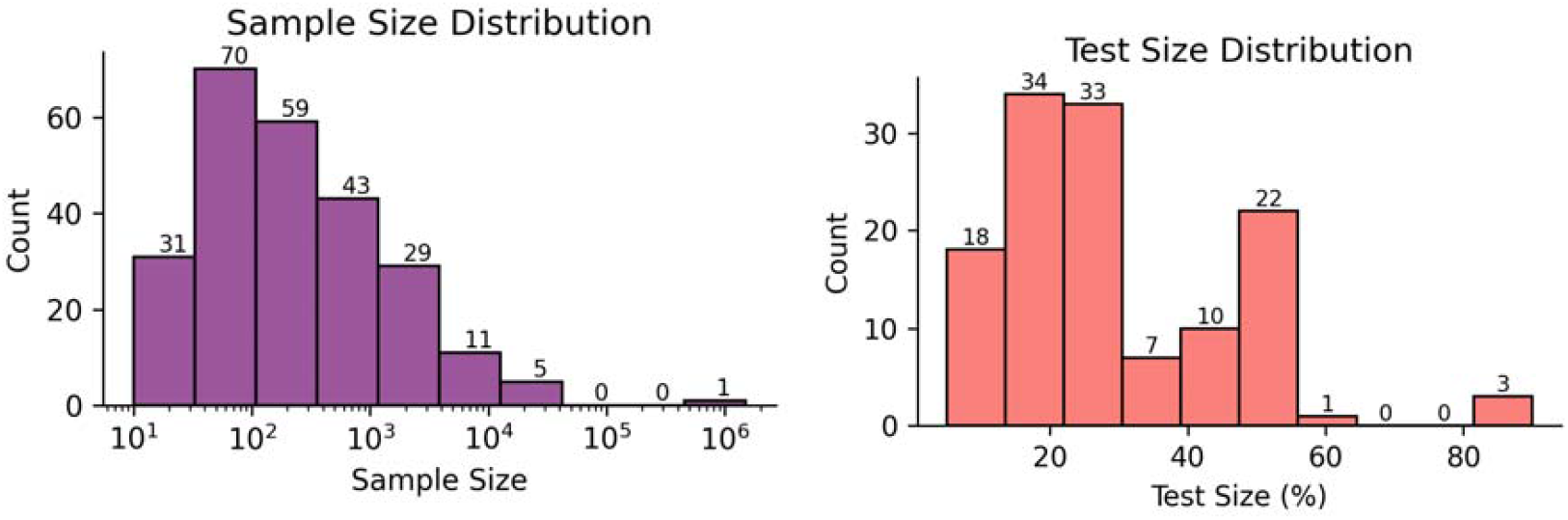
Dataset size and test-set representation. (a) Distribution of Sample Size (log scale) and (b) the proportion of data separated for test.

Our findings reveal that some studies employ robust validation methods, such as 10-fold cross-validation in 52 studies (20%) and 5-fold cross-validation used in 33 studies (12%). Also, 28 studies (11%) reported using the leave-one-out cross-validation technique; this occurred most frequently in smaller datasets or studies where maximum utilization of available data was critical.

## Discussion

Our findings highlight the diverse applications of machine learning OSA research. We found a with primary focus on applications related to improving diagnostic methods (29%), identification of apneas and hypopneas at the epoch level (11%), and snoring detection (10%). This trend is likely driven by the need for automatic diagnosis tools as an alternative to the high costs of complex PSG procedures, which require patients to stay overnight in sleep laboratories. Moreover, there is a strong emphasis on replicating or enhancing AHI-based approaches for diagnosis, similar to recent alternative metrics such as hypoxic burden, which may better reflect the physiological impact of OSA and its association with cardiovascular outcomes [19]. However, within the timeframe of our review, we found that such emerging metrics were rarely incorporated into machine learning studies, likely due to their recent development and limited integration into standard datasets. Future work has the potential to combine machine learning and newer alternative OSA-related metrics, possibly combining them, seeking more informative risk stratification capabilities.

The emergence of innovative sensors in wearable devices and advancements in machine learning techniques further underscore the interest in developing more efficient diagnostic methodologies [20]. An interesting example is the work of Gu et al. (2020), who used a novel pulse oximeter combined with an accelerometer and a trained neural network to compute the Apnea-Hypopnea Index (AHI), achieving a Pearson correlation score of 0.9, where 1 is a perfect correlation [21]. Despite these promising developments, our review shows that only a small proportion of machine learning applications in OSA research focus on treatment-related aspects. Specifically, studies involving wearable or nearable devices constitute just 6% of the total (n=15), and those addressing treatment and surgical candidacy represent less than 5% (n=13). This disparity underscores a substantial opportunity for future research to expand beyond diagnosis and explore machine learning’s role in optimizing treatment strategies and long-term patient management.

In our demographic analysis, we were interested in identifying the different representations of population subgroups, shedding light on the heterogeneity of the current research landscape. Considering BMI, most of the populations in the studies were overweight but not severely obese. The age distribution shows a bimodal pattern, capturing both pediatric participants and adults in the 40-60-year age range, which is a critical period for the emergence of OSA risk factors and symptoms [22]. We identified significant gaps in research, specifically the low representation of studies involving younger obese adults and populations older than 60 years. The growing number of older individuals in most developed countries, coupled with the higher prevalence of OSA in these groups (36%), highlights a critical gap in the literature[23].

Our sex-specific analysis highlighted a significant imbalance between males and females in datasets used to train machine learning models. This underscores the need to be attentive to gender distribution during the data collection process, as well as including more women in samples to better represent sex as a biological variable in clinical research. Consistent with recent discussions on bias mitigation strategies in healthcare AI, our results suggest the need for clear methodological guidelines addressing demographic representation. These guidelines should focus not only on achieving statistical balance but also on understanding the structural and organizational contexts in which data are collected and used [24].

Geographically, the studies are well-distributed across continents. Still, the lack of data on reported race and ethnicity reveals a potential gap in inclusivity, with a small number of studies explicitly including multiracial or multiethnic samples. This demographic overview highlights the need for more balanced and inclusive research approaches to better address OSA across different populations. Moreover, this finding hits the current core demand for a discussion on machine learning ethics and how careful consideration is needed in terms of ethical guidelines to ensure equitable and unbiased models [25].

We found that 83 studies (32%) utilized multiple data sources, underscoring the complexity of data integration in OSA research, where multiple sources of physiological data highlight the need for a multi-modal approach in machine learning. Integrating data from diverse physiological signals, clinical records, questionnaires, and wearable devices significantly enriches the feature set available for machine learning models, potentially enhancing their predictive power. However, such integration may complicate the preprocessing, feature extraction, and ultimately the interpretability of model outcomes [26].

Electrocardiogram (ECG) was the most frequently used data type, appearing in 27% of studies, often sourced from open-access datasets such as Physionet’s Apnea ECG dataset [27]. Another example, Holfinger et al. (2022), used the Sleep Heart Health Study, a large cohort (N=5,599) publicly available at the National Sleep Research Resource (https://sleepdata.org/), to evaluate machine learning models for diagnosing OSA. The growing reliance on these public datasets highlights efforts to address healthcare data scarcity, which commonly arises from patient privacy concerns and fragmented healthcare data infrastructures [28].

The frequent use of specific machine learning techniques observed in our review aligns with current research trends, particularly the rising interest in deep learning models. Additionally, we noted substantial usage of structured tabular data, including demographics and questionnaire results, paired with support vector machines (SVM) and ensemble models, such as Random Forest and XGBoost, representing current state-of-the-art approaches for this data type. Although test set proportions (around 20% of the total data) and validation techniques (commonly 5-or 10-fold cross-validation) adhered to standard practice, we observed that more than half of the studies employed small sample sizes (fewer than 1,000 samples), underscoring persistent challenges related to data availability and acquisition.

In terms of evaluation metrics for classification, we observed that most studies reported values around 80%, independent of the metric, with some studies achieving 100% for certain metrics. True perfect scores are extremely rare, especially in large datasets, which raises two significant concerns: 1) the possibility of inflated scores and 2) the risk of overfitting machine learning models. In the case of overfitting, the model’s performance is not reproducible outside its own research [29].

In contrast to other reviews on machine learning applied to OSA research, our analysis reveals a broader landscape regarding data heterogeneity, demographic bias, and OSA-specific topics beyond diagnosis [13]. Still, it mirrors similar concerns regarding model development practice. Such contrast emphasizes the continued need for stronger methodological standards and reproducibility practices, especially concerning demographic diversity, sample size, and multimodal applications, to improve generalizability and fairness in OSA-related machine learning research.

The scope of this narrative review was limited to studies published between January 2018 and March 2023, aligning with our objective to evaluate the use of machine learning in OSA research with a focus on traditional methods, such as feature-based algorithms and classic neural networks. This period occurred before the widespread adoption of large language models (LLMs) and foundation models [30]. In particular, March 2023 marked an extraordinary moment in the LLM landscape with the release of OpenAI’s GPT-4 [31]. Although these advanced models rapidly influence medical applications, their use remains in early, exploratory stages [32]. Still, we anticipate a transition from traditional machine learning techniques toward LLM and foundation model-based approaches post-2023, enabling methods that harness the largely available text and unstructured multimodal data inherent to polysomnograms. Ultimately, our review provides a necessary baseline for comparison with future studies capturing this emerging shift.

We acknowledge that our work is also limited given the use of the PubMed database during the search process, which may have excluded studies from databases like Scopus, Web of Science, or IEEE. PubMed was chosen for its peer-reviewed content and relevance to our clinical focus. While broader inclusion could have enriched the dataset, it would have required significantly more data curation and management resources. Also, as a future direction, we propose a deeper quantitative analysis and text mining to get insights from titles and content [33], as well as conduct topic modeling to uncover latent themes and trends across the corpus [34].

## Recommendations

Overall, our findings indicated that trained models in OSA research are often underrepresented in women, younger obese adults, individuals over 60 years old, and diverse racial groups. Additionally, we found a high prevalence of small sample sizes for training models, highlighting the challenges in healthcare data acquisition. Considering the findings of this work and seeking to guide future research, focusing on transparent, robust, and ethical deployment of machine learning for OSA diagnosis and management, we offer the following recommendations for future researchers:

### Sample Size

Unlike traditional statistical aproaches, where sample sizes can be derived based on effect sizes, machine learning applications require sample size considerations that depend on the specific context, prediction method, number of features, and desired predictive performance. Having a preliminary dataset available can help estimate an appropriate total sample size, which is often related to clinical impact. For external evaluation, it is suggested that the sample for external validation should have at least 100 events per outcome [35].

### Validation Strategies

Prioritize external validation using independent datasets to assess model generalizability. At a minimum, internal validation with clearly documented cross-validation techniques (e.g., stratified k-fold) should be reported [36]. We also recommend including confidence intervals when performing cross-validation and a comprehensive set of performance metrics beyond accuracy (e.g., sensitivity, specificity, F1-Score, area under the ROC curve, area under the precision-recall curve, and confusion matrix).

### Multimodal Integration

Leverage the potential of multimodal data, including physiological signals, clinical records, wearable devices, images, and patient-reported outcomes, to improve predictive performance. Newer deep learning models have shown great ability to fuse information from different data types. However, obtaining multimodal datasets of significant sizes is challenging due to regulatory complexities, limited collaboration between those who manage different databases, and even technical complications such as the alignment of longitudinal data [26].

### Demographic Balance

Intentionally design studies to include underrepresented populations to improve model fairness and equity in clinical application. Note that having a completely unbiased, balanced dataset is almost impossible. Thus, distinguish between desirable and undesirable biases and be transparent about them when reporting analysis. Implement explainable algorithms to provide transparent interpretation of the model outputs, which detect potential algorithm misbehavior towards specific subpopulations [37]. Bias correction may also be implemented via training models on specific demographic groups, avoiding the one-model-predicts-all convention. However, this approach does not guarantee overall optimal performance [38]. Utilizing demographic balancing strategies during data collection and model training phases can further mitigate bias [24].

## Conclusion

We provided a narrative review of 254 scientific articles published between January 2018 and March 2023 that utilized machine learning in obstructive sleep apnea (OSA) research. Key trends identified include: 1) a predominant focus on classification tasks, leveraging mainly deep learning models and support vector machines; 2) studied cohorts were primarily overweight males, with significant underrepresentation of women, younger obese adults, individuals older than 60 years, and diverse racial groups; 3) frequent utilization of multiple physiological data sources, especially ECG signals, often obtained from public datasets; and 4) prevalent use of small sample sizes (fewer than 1,000 participants) and limited application of robust validation methodologies, reflecting ongoing challenges in healthcare data acquisition. Additionally, our review highlights gaps in treatment-focused studies and the limited incorporation of emerging OSA-related metrics beyond traditional AHI scoring. To help advance the field, we provided a set of recommendations addressing the main pitfalls, including sample size, validation strategies, multimodal integration, and demographic balancing to enhance the reliability, transparency, and generalizability of future machine learning research in OSA. Future work may also cover aspects such as model interpretability, clinical workflow integration, or regulatory and other ethical considerations.

## Data Availability

All data produced in the present study are available upon reasonable request to the authors

## Summary Table

- The review analyzed 254 studies published between 2018 and 2023, finding that most applied machine learning to classify and diagnose obstructive sleep apnea (OSA), with deep learning models being the most popular techniques used.
- There are severe demographic gaps where most study populations were predominantly overweight males and an underrepresentation of women, younger obese adults, individuals over 60 years old, and diverse racial groups, indicating a need for more inclusive research to improve generalizability. Multiple data sources, such as polysomnography, wearables, and electrocardiograms, were used, but data collection methods lacked standardization. Many studies had small sample sizes and limited use of robust machine learning validation strategies, potentially affecting the reliability of machine learning applications.
- We provide recommendations that address the choice of sample sizes, validation strategies, multimodal data integration, and demographic balancing to enhance the reliability, transparency, and generalizability of future machine learning research in OSA.

## Acknowledgments

The research reported in this publication was supported by the National Heart Lung and Blood Institute of the National Institutes of Health under award number 1R21HL170206-01. We also acknowledge Dr. John Fredieu for the manuscript writing review and Mary Schleicher and Dr. Christian Mouchati for their contributions to the initial data collection.

## Author contributions: CRediT

Matheus Lima Diniz Araujo, Trevor Winger, Louis Kazaglis, Piyush Mathur, and Reena Mehra were involved in the conception and methodology. Reena Mehra contributed with supervision. Matheus Lima Diniz Araujo and Trevor Winger contributed to data curation, formal analysis, visualization, and original draft writing. Samer Ghosn, Carl Saab, and Jaideep Srivastava contributed to subsequent draft revisions.

### Ethics

Not relevant due to the nature of this manuscript.

### Statement on conflicts of interest

All authors declare no competing financial or non-financial interests.

## Footnotes

1 Available www.covidence.org

## Notes

### Competing Interest Statement

The authors have declared no competing interest.

### Funding Statement

The research reported in this publication was partially supported by the National Heart Lung and Blood Institute of the National Institutes of Health under award number 1R21HL170206-01.

### Summary of Updates

Abstract and Introduction clarified. Discussion improved. This updates the manuscript after peer reviewed process.

