## Supplemental Materials for "Status and Opportunities of Machine Learning Applications in Obstructive Sleep Apnea: A Narrative Review"

### Supplementary Material:

| Title | Authors | Published Year | Published Month | Journal | Volume | Issue | Pages | Accession Number | DOI |
| --- | --- | --- | --- | --- | --- | --- | --- | --- | --- |
| A 2D convolutional neural network to detect sleep apnea in children using airflow and oximetry. | JimÃ©nez-GarcÃ­a J; GarcÃ­a M; GutiÃ©rrez-Tobal GC; Kheirandish-Gozal L; Vaquerizo-Villar F; Ãlvarez D; Del Campo F; Gozal D; Hornero R | 2022 | Aug | Comput Biol Med | 147 |  | 105784 |  | 10.1016/j.compbiomed.2022.105784 |
| A Bag of Wavelet Features for Snore Sound Classification. | Qian K; Schmitt M; Janott C; Zhang Z; Heiser C; Hohenhorst W; Herzog M; Hemmert W; Schuller B | 2019 | Apr | Ann Biomed Eng | 47 | 4 | 1000-1011 | | 10.1007/s10439-019-02217-0 |
| A Classifying Model of Obstructive Sleep Apnea Based on Heart Rate Variability in a Large Korean Population. | Park P; Kim JW | 2023 | Feb | J Korean Med Sci | 38 | 7 | e49 |  | 10.3346/jkms.2023.38.e49 |
| A comparison of probabilistic classifiers for sleep stage classification. | Fonseca P; den Teuling N; Long X; Aarts RM | 2018 | May | Physiol Meas | 39 | 5 | 55001 |  | 10.1088/1361-6579/aabbc2 |
| A comparison of regularized logistic regression and random forest machine learning models for daytime diagnosis of obstructive sleep apnea. | Hajipour F; Jozani MJ; Moussavi Z | 2020 | Oct | Med Biol Eng Comput | 58 | 10 | 2517-2529 | | 10.1007/s11517-020-02206-9 |
| A Comparison of Signal Combinations for Deep Learning-Based Simultaneous Sleep Staging and Respiratory Event Detection. | Huttunen R; Leppanen T; Duce B; Arnardottir ES; Nikkonen S; Myllymaa S; Toyras J; Korkalainen H | 2022 | Nov | IEEE Trans Biomed Eng | PP |  |  |  | 10.1109/TBME.2022.3225268 |
| A Convolutional Neural Network Architecture to Enhance Oximetry Ability to Diagnose Pediatric Obstructive Sleep Apnea. | Vaquerizo-Villar F; Alvarez D; Kheirandish-Gozal L; Gutierrez-Tobal GC; Barroso-Garcia V; Santamaria-Vazquez E; Campo FD; Gozal D; Hornero R | 2021 | Aug | IEEE J Biomed Health Inform | 25 | 8 | 2906-2916 | | 10.1109/JBHI.2020.3048901 |
| A deep learning-based decision support system for diagnosis of OSAS using PTT signals. | Arslan Tuncer S; AkÄ±lotu B; Toraman S | 2019 | Jun | Med Hypotheses | 127 |  | 15-22 |  | 10.1016/j.mehy.2019.03.026 |
| A fused-image-based approach to detect obstructive sleep apnea using a single-lead ECG and a 2D convolutional neural network. | Niroshana SMI; Zhu X; Nakamura K; Chen W | 2021 |  | PLoS One | 16 | 4 | e0250618 |  | 10.1371/journal.pone.0250618 |
| A Fuzzy Neural Network Model for Rapid Prediction of Optimal Positive Airway Pressures in OSAS Patients. | Juang CF; Pan GR; Wen CY; Chang KM; Wu MF; Huang WC | 2022 | Apr | IEEE J Biomed Health Inform | 26 | 4 | 1506-1515 | | 10.1109/JBHI.2021.3120662 |
| A machine learning-based test for adult sleep apnoea screening at home using oximetry and airflow. | Ãlvarez D; Cerezo-HernÃ¡ndez A; Crespo A; GutiÃ©rrez-Tobal GC; Vaquerizo-Villar F; Barroso-GarcÃ­a V; Moreno F; Arroyo CA; Ruiz T; Hornero R; Del Campo F | 2020 | Mar | Sci Rep | 10 | 1 | 5332 |  | 10.1038/s41598-020-62223-4 |
| A model for obstructive sleep apnea detection using a multi-layer feed-forward neural network based on electrocardiogram, pulse oxygen saturation, and body mass index. | Li Z; Li Y; Zhao G; Zhang X; Xu W; Han D | 2021 | Dec | Sleep Breath | 25 | 4 | 2065-2072 | | 10.1007/s11325-021-02302-6 |
| A New Berlin Questionnaire Simplified by Machine Learning Techniques in a Population of Italian Healthcare Workers to Highlight the Suspicion of Obstructive Sleep Apnea. | De Nunzio G; Conte L; Lupo R; Vitale E; CalabrÃ² A; Ercolani M; Carvello M; Arigliani M; Toraldo DM; De Benedetto L | 2022 |  | Front Med (Lausanne) | 9 |  | 866822 |  | 10.3389/fmed.2022.866822 |
| A New Feature with the Potential to Detect the Severity of Obstructive Sleep Apnoea via Snoring Sound Analysis. | Hayashi S; Tamaoka M; Tateishi T; Murota Y; Handa I; Miyazaki Y | 2020 | Apr | Int J Environ Res Public Health | 17 | 8 |  |  | 10.3390/ijerph17082951 |
| A Novel Clinical Method for Detecting Obstructive Sleep Apnea using of Nonlinear Mapping. | Karimi Moridani M | 2022 | Feb | J Biomed Phys Eng | 12 | 1 | 31-34 |  | 10.31661/jbpe.v0i0.1211 |
| A Novel Decision Making Procedure during Wakefulness for Screening Obstructive Sleep Apnea using Anthropometric Information and Tracheal Breathing Sounds. | Elwali A; Moussavi Z | 2019 | Aug | Sci Rep | 9 | 1 | 11467 |  | 10.1038/s41598-019-47998-5 |
| A novel deep domain adaptation method for automated detection of sleep apnea/hypopnea events. | Du Z; Wang J; Ren Y | 2023 | Feb | Physiol Meas | 44 | 1 |  |  | 10.1088/1361-6579/aca879 |
| A novel deep feature transfer-based OSA detection method using sleep sound signals. | Luo J; Liu H; Gao X; Wang B; Zhu X; Shi Y; Hei X; Ren X | 2020 | Aug | Physiol Meas | 41 | 7 | 75009 |  | 10.1088/1361-6579/ab9e7b |
| A Novel Model to Estimate Key Obstructive Sleep Apnea Endotypes from Standard Polysomnography and Clinical Data and Their Contribution to Obstructive Sleep Apnea Severity. | Dutta R; Delaney G; Toson B; Jordan AS; White DP; Wellman A; Eckert DJ | 2021 | Apr | Ann Am Thorac Soc | 18 | 4 | 656-667 |  | 10.1513/AnnalsATS.202001-064OC |
| A novel, simple, and accurate pulse oximetry indicator for screening adult obstructive sleep apnea. | Nigro CA; CastaÃ±o G; Bledel I; Colombi A; Zicari MC | 2022 | Sep | Sleep Breath | 26 | 3 | 1125-1134 | | 10.1007/s11325-021-02439-4 |
| A Sleep Apnea Detection System Based on a One-Dimensional Deep Convolution Neural Network Model Using Single-Lead Electrocardiogram. | Chang HY; Yeh CY; Lee CT; Lin CC | 2020 | Jul | Sensors (Basel) | 20 | 15 |  |  | 10.3390/s20154157 |
| Accelerometry-derived respiratory index estimating apnea-hypopnea index for sleep apnea screening. | Bricout A; Fontecave-Jallon J; PÃ©pin JL; GumÃ©ry PY | 2021 | Aug | Comput Methods Programs Biomed | 207 |  | 106209 |  | 10.1016/j.cmpb.2021.106209 |
| Accurate Deep Learning-Based Sleep Staging in a Clinical Population With Suspected Obstructive Sleep Apnea. | Korkalainen H; Aakko J; Nikkonen S; Kainulainen S; Leino A; Duce B; Afara IO; Myllymaa S; Toyras J; Leppanen T | 2020 | Jul | IEEE J Biomed Health Inform | 24 | 7 | 2073-2081 | | 10.1109/JBHI.2019.2951346 |
| Achieving Accurate Automatic Sleep Apnea/Hypopnea Syndrome Assessment Using Nasal Pressure Signal. | Lin YS; Wu YP; Wu YC; Lee PL; Yang CH | 2022 | Nov | IEEE J Biomed Health Inform | 26 | 11 | 5473-5481 | | 10.1109/JBHI.2022.3199454 |
| Acoustic Screening for Obstructive Sleep Apnea in Home Environments Based on Deep Neural Networks. | Romero HE; Ma N; Brown GJ; Hill EA | 2022 | Jul | IEEE J Biomed Health Inform | 26 | 7 | 2941-2950 | | 10.1109/JBHI.2022.3154719 |
| Advanced Proteomics and Cluster Analysis for Identifying Novel Obstructive Sleep Apnea Subtypes before and after CPAP Therapy. | Kundel V; Cohen O; Khan S; Patel M; Kim-Schulze S; Kovacic J; SuÃ¡rez-FariÃ±as M; Shah NA | 2023 | Feb | Ann Am Thorac Soc | |  |  |  | 10.1513/AnnalsATS.202210-897OC |
| AI-based automatic segmentation of craniomaxillofacial anatomy from CBCT scans for automatic detection of pharyngeal airway evaluations in OSA patients. | Orhan K; Shamshiev M; Ezhov M; Plaksin A; Kurbanova A; Ãœnsal G; Gusarev M; Golitsyna M; Aksoy S; MÄ±sÄ±rlÄ± M; Rasmussen F; Shumilov E; Sanders A | 2022 | Jul | Sci Rep | 12 | 1 | 11863 |  | 10.1038/s41598-022-15920-1 |
| AIOSA: An approach to the automatic identification of obstructive sleep apnea events based on deep learning. | Bernardini A; Brunello A; Gigli GL; Montanari A; Saccomanno N | 2021 | Aug | Artif Intell Med | 118 |  | 102133 |  | 10.1016/j.artmed.2021.102133 |
| Amplitude spectrum trend-based feature for excitation location classification from snore sounds. | Sun J; Hu X; Chen C; Peng S; Ma Y | 2020 | Sep | Physiol Meas | 41 | 8 | 85006 |  | 10.1088/1361-6579/abaa34 |
| An Empirical Study of Questionnaires for the Diagnosis of Pediatric Obstructive Sleep Apnea. | Ahmed S; Hasani S; Koone M; Thirumuruganathan S; Diaz-Abad M; Mitchell R; Isaiah A; Das G | 2018 | Jul | Annu Int Conf IEEE Eng Med Biol Soc | 2018 |  | 4097-4100 | | 10.1109/EMBC.2018.8513389 |
| An open-source, high-performance tool for automated sleep staging. | Vallat R; Walker MP | 2021 | Oct | Elife | 10 |  |  |  | 10.7554/eLife.70092 |
| An OSAHS evaluation method based on multi-features acoustic analysis of snoring sounds. | Jiang Y; Peng J; Song L | 2021 | Aug | Sleep Med | 84 |  | 317-323 |  | 10.1016/j.sleep.2021.06.012 |
| Apnea and Hypopnea Events Classification Using Amplitude Spectrum Trend Feature of Snores. | Sun J; Hu X; Zhao Y; Sun S; Chen C; Peng S | 2018 | Jul | Annu Int Conf IEEE Eng Med Biol Soc | 2018 |  | 6036-6039 | | 10.1109/EMBC.2018.8513688 |
| Application of an optimal class of antisymmetric wavelet filter banks for obstructive sleep apnea diagnosis using ECG signals. | Sharma M; Agarwal S; Acharya UR | 2018 | Sep | Comput Biol Med | 100 |  | 100-113 |  | 10.1016/j.compbiomed.2018.06.011 |
| Application of machine learning to predict obstructive sleep apnea syndrome severity. | Mencar C; Gallo C; Mantero M; Tarsia P; Carpagnano GE; Foschino Barbaro MP; Lacedonia D | 2020 | Mar | Health Informatics J | 26 | 1 | 298-317 |  | 10.1177/1460458218824725 |
| Artificial Intelligence Analysis of Mandibular Movements Enables Accurate Detection of Phasic Sleep Bruxism in OSA Patients: A Pilot Study. | Martinot JB; Le-Dong NN; Cuthbert V; Denison S; Gozal D; Lavigne G; PÃ©pin JL | 2021 |  | Nat Sci Sleep | 13 |  | 1449-1459 | | 10.2147/NSS.S320664 |
| Artificial neural network analysis of the oxygen saturation signal enables accurate diagnostics of sleep apnea. | Nikkonen S; Afara IO; LeppÃ¤nen T; TÃ¶yrÃ¤s J | 2019 | Sep | Sci Rep | 9 | 1 | 13200 |  | 10.1038/s41598-019-49330-7 |
| Assessment of Mandibular Movement Monitoring With Machine Learning Analysis for the Diagnosis of Obstructive Sleep Apnea. | PÃ©pin JL; Letesson C; Le-Dong NN; Dedave A; Denison S; Cuthbert V; Martinot JB; Gozal D | 2020 | Jan | JAMA Netw Open | 3 | 1 | e1919657 |  | 10.1001/jamanetworkopen.2019.19657 |
| Assessment of obstructive sleep apnea-related sleep fragmentation utilizing deep learning-based sleep staging from photoplethysmography. | Huttunen R; LeppÃ¤nen T; Duce B; Oksenberg A; Myllymaa S; TÃ¶yrÃ¤s J; Korkalainen H | 2021 | Oct | Sleep | 44 | 10 |  |  | 10.1093/sleep/zsab142 |
| Association between physical activity and risk of obstructive sleep apnea. | Duan X; Zheng M; He S; Lao L; Huang J; Zhao W; Lao XQ; Deng H; Liu X | 2021 | Dec | Sleep Breath | 25 | 4 | 1925-1934 | | 10.1007/s11325-021-02318-y |
| Association of hypoglossal nerve stimulator response with machine learning identified negative effort dependence patterns. | Lou B; Rusk S; Nygate YN; Quintero L; Ishikawa O; Shikowitz M; Greenberg H | 2022 | May | Sleep Breath | |  | 7-Jan |  | 10.1007/s11325-022-02641-y |
| Association of snoring characteristics with predominant site of collapse of upper airway in obstructive sleep apnea patients. | Sebastian A; Cistulli PA; Cohen G; de Chazal P | 2021 | Dec | Sleep | 44 | 12 |  |  | 10.1093/sleep/zsab176 |
| At-home wireless monitoring of acute hemodynamic disturbances to detect sleep apnea and sleep stages via a soft sternal patch. | Zavanelli N; Kim H; Kim J; Herbert R; Mahmood M; Kim YS; Kwon S; Bolus NB; Torstrick FB; Lee CSD; Yeo WH | 2021 | Dec | Sci Adv | 7 | 52 | eabl4146 |  | 10.1126/sciadv.abl4146 |
| Audio-based snore detection using deep neural networks. | Xie J; Aubert X; Long X; van Dijk J; Arsenali B; Fonseca P; Overeem S | 2021 | Mar | Comput Methods Programs Biomed | 200 |  | 105917 |  | 10.1016/j.cmpb.2020.105917 |
| Auditory Receptive Field Net based Automatic Snore Detection for Wearable Devices. | Hu X; Sun J; Dong J; Zhang X | 2022 | Apr | IEEE J Biomed Health Inform | PP |  |  |  | 10.1109/JBHI.2022.3164517 |
| Automated Detection of Obstructive Sleep Apnea Events from a Single-Lead Electrocardiogram Using a Convolutional Neural Network. | Urtnasan E; Park JU; Joo EY; Lee KJ | 2018 | Apr | J Med Syst | 42 | 6 | 104 |  | 10.1007/s10916-018-0963-0 |
| Automated Detection of Sleep Apnea-Hypopnea Events Based on 60 GHz Frequency-Modulated Continuous-Wave Radar Using Convolutional Recurrent Neural Networks: A Preliminary Report of a Prospective Cohort Study. | Choi JW; Kim DH; Koo DL; Park Y; Nam H; Lee JH; Kim HJ; Hong SN; Jang G; Lim S; Kim B | 2022 | Sep | Sensors (Basel) | 22 | 19 |  |  | 10.3390/s22197177 |
| Automated identification of the predominant site of upper airway collapse in obstructive sleep apnoea patients using snore signal. | Sebastian A; Cistulli PA; Cohen G; de Chazal P | 2020 | Oct | Physiol Meas | 41 | 9 | 95005 |  | 10.1088/1361-6579/abaa33 |
| Automated Scoring of Respiratory Events in Sleep With a Single Effort Belt and Deep Neural Networks. | Nassi TE; Ganglberger W; Sun H; Bucklin AA; Biswal S; van Putten MJAM; Thomas RJ; Westover MB | 2022 | Jun | IEEE Trans Biomed Eng | 69 | 6 | 2094-2104 | | 10.1109/TBME.2021.3136753 |
| Automatic classification of excitation location of snoring sounds. | Sun J; Hu X; Peng S; Peng CK; Ma Y | 2021 | May | J Clin Sleep Med | 17 | 5 | 1031-1038 | | 10.5664/jcsm.9094 |
| Automatic classification of the obstruction site in obstructive sleep apnea based on snoring sounds. | Liu Y; Feng Y; Li Y; Xu W; Wang X; Han D | 2022 | Nov-Dec | Am J Otolaryngol | 43 | 6 | 103584 |  | 10.1016/j.amjoto.2022.103584 |
| Automatic Detection of Obstructive Sleep Apnea Events Using a Deep CNN-LSTM Model. | Zhang J; Tang Z; Gao J; Lin L; Liu Z; Wu H; Liu F; Yao R | 2021 |  | Comput Intell Neurosci | 2021 |  | 5594733 |  | 10.1155/2021/5594733 |
| Automatic detection of respiratory events during sleep from Polysomnography data using Layered Hidden Markov Model. | Sadoughi A; Shamsollahi MB; Fatemizadeh E | 2022 | Jan | Physiol Meas | 43 | 1 |  |  | 10.1088/1361-6579/ac45e1 |
| Automatic identification of respiratory events based on nasal airflow and respiratory effort of the chest and abdomen. | Liu J; Li Q; Chen Y; Wang B; Li Y; Xin Y | 2021 | Jul | Physiol Meas | 42 | 7 |  |  | 10.1088/1361-6579/abfae5 |
| Automatic Respiratory Event Scoring in Obstructive Sleep Apnea Using a Long Short-Term Memory Neural Network. | Nikkonen S; Korkalainen H; Leino A; Myllymaa S; Duce B; Leppanen T; Toyras J | 2021 | Aug | IEEE J Biomed Health Inform | 25 | 8 | 2917-2927 | | 10.1109/JBHI.2021.3064694 |
| Automatic scoring of drug-induced sleep endoscopy for obstructive sleep apnea using deep learning. | Hanif U; Kiaer EK; Capasso R; Liu SY; Mignot EJM; Sorensen HBD; Jennum P | 2023 | Feb | Sleep Med | 102 |  | 19-29 |  | 10.1016/j.sleep.2022.12.015 |
| Automatic Segmentation and QuantificationÂ of Upper Airway Anatomic Risk Factors for Obstructive Sleep Apnea on Unprocessed Magnetic Resonance Images. | Bommineni VL; Erus G; Doshi J; Singh A; Keenan BT; Schwab RJ; Wiemken A; Davatzikos C | 2023 | Mar | Acad Radiol | 30 | 3 | 421-430 |  | 10.1016/j.acra.2022.04.023 |
| Automatic segmentation of the pharyngeal airway space with convolutional neural network. | Shujaat S; Jazil O; Willems H; Van Gerven A; Shaheen E; Politis C; Jacobs R | 2021 | Aug | J Dent | 111 |  | 103705 |  | 10.1016/j.jdent.2021.103705 |
| Automatic snoring sounds detection from sleep sounds based on deep learning. | Jiang Y; Peng J; Zhang X | 2020 | Jun | Phys Eng Sci Med | 43 | 2 | 679-689 |  | 10.1007/s13246-020-00876-1 |
| BASH-GN: a new machine learning-derived questionnaire for screening obstructive sleep apnea. | Huo J; Quan SF; Roveda J; Li A | 2022 | Apr | Sleep Breath | |  |  |  | 10.1007/s11325-022-02629-8 |
| Belun Ring Platform: a novel home sleep apnea testing system for assessment of obstructive sleep apnea. | Gu W; Leung L; Kwok KC; Wu IC; Folz RJ; Chiang AA | 2020 | Sep | J Clin Sleep Med | 16 | 9 | 1611-1617 | ClinicalTrials.gov/NCT04121923 | 10.5664/jcsm.8592 |
| Bispectral Analysis of Heart Rate Variability to Characterize and Help Diagnose Pediatric Sleep Apnea. | MartÃ­n-Montero A; GutiÃ©rrez-Tobal GC; Gozal D; Barroso-GarcÃ­a V; Ãlvarez D; Del Campo F; Kheirandish-Gozal L; Hornero R | 2021 | Aug | Entropy (Basel) | 23 | 8 |  |  | 10.3390/e23081016 |
| Blood-based lipidomic signature of severe obstructive sleep apnoea in Alzheimer's disease. | Dakterzada F; BenÃ­tez ID; Targa A; Carnes A; Pujol M; JovÃ© M; MÃ­nguez O; Vaca R; SÃ¡nchez-de-la-Torre M; BarbÃ© F; Pamplona R; PiÃ±ol-Ripoll G | 2022 | Nov | Alzheimers Res Ther | 14 | 1 | 163 | ClinicalTrials.gov/NCT02814045 | 10.1186/s13195-022-01102-8 |
| Brief digital sleep questionnaire powered by machine learning prediction models identifies common sleep disorders. | Schwartz AR; Cohen-Zion M; Pham LV; Gal A; Sowho M; Sgambati FP; Klopfer T; Guzman MA; Hawks EM; Etzioni T; Glasner L; Druckman E; Pillar G | 2020 | Jul | Sleep Med | 71 |  | 66-76 |  | 10.1016/j.sleep.2020.03.005 |
| Capacitively-Coupled ECG and Respiration for Sleep-Wake Prediction and Risk Detection in Sleep Apnea Patients. | Huysmans D; Castro I; BorzÃ©e P; Patel A; Torfs T; Buyse B; Testelmans D; Van Huffel S; Varon C | 2021 | Sep | Sensors (Basel) | 21 | 19 |  |  | 10.3390/s21196409 |
| Cardiovascular risk and mortality prediction in patients suspected of sleep apnea: a model based on an artificial intelligence system. | Blanchard M; Feuilloy M; GervÃ¨s-PinquiÃ© C; Trzepizur W; Meslier N; Goupil F; Pigeanne T; Racineux JL; Balusson F; Oger E; Gagnadoux F; Girault JM | 2021 | Oct | Physiol Meas | 42 | 10 |  |  | 10.1088/1361-6579/ac2a8f |
| Central apnea detection in premature infants using machine learning. | Varisco G; Peng Z; Kommers D; Zhan Z; Cottaar W; Andriessen P; Long X; van Pul C | 2022 | Nov | Comput Methods Programs Biomed | 226 |  | 107155 |  | 10.1016/j.cmpb.2022.107155 |
| Characteristics of salivary microbiota in children with obstructive sleep apnea: A prospective study with polysomnography. | Huang X; Chen X; Gong X; Xu Y; Xu Z; Gao X | 2022 |  | Front Cell Infect Microbiol | 12 |  | 945284 |  | 10.3389/fcimb.2022.945284 |
| Classification and Detection of Breathing Patterns with Wearable Sensors and Deep Learning. | McClure K; Erdreich B; Bates JHT; McGinnis RS; Masquelin A; Wshah S | 2020 | Nov | Sensors (Basel) | 20 | 22 |  |  | 10.3390/s20226481 |
| Classification of Pharynx from MRI Using a Visual Analysis Tool to Study Obstructive Sleep Apnea. | Shahid MLUR; Mir J; Shaukat F; Saleem MK; Tariq MAUR; Nouman A | 2021 |  | Curr Med Imaging | 17 | 5 | 613-622 |  | 10.2174/1573405616666201118143935 |
| Classification of severe obstructive sleep apnea with cognitive impairment using degree centrality: A machine learning analysis. | Liu X; Shu Y; Yu P; Li H; Duan W; Wei Z; Li K; Xie W; Zeng Y; Peng D | 2022 |  | Front Neurol | 13 |  | 1005650 |  | 10.3389/fneur.2022.1005650 |
| Classification of sleep apnea based on EEG sub-band signal characteristics. | Zhao X; Wang X; Yang T; Ji S; Wang H; Wang J; Wang Y; Wu Q | 2021 | Mar | Sci Rep | 11 | 1 | 5824 |  | 10.1038/s41598-021-85138-0 |
| Classification of Sleep Apnea Severity by Electrocardiogram Monitoring Using a Novel Wearable Device. | Baty F; Boesch M; Widmer S; Annaheim S; Fontana P; Camenzind M; Rossi RM; Schoch OD; Brutsche MH | 2020 | Jan | Sensors (Basel) | 20 | 1 |  |  | 10.3390/s20010286 |
| Clinical validation of a mandibular movement signal based system for the diagnosis of pediatric sleep apnea. | Martinot JB; Cuthbert V; Le-Dong NN; Coumans N; De Marneffe D; Letesson C; PÃ©pin JL; Gozal D | 2022 | Aug | Pediatr Pulmonol | 57 | 8 | 1904-1913 | | 10.1002/ppul.25320 |
| Cloud algorithm-driven oximetry-based diagnosis of obstructive sleep apnoea in symptomatic habitually snoring children. | Xu Z; GutiÃ©rrez-Tobal GC; Wu Y; Kheirandish-Gozal L; Ni X; Hornero R; Gozal D | 2019 | Feb | Eur Respir J | 53 | 2 |  |  | 10.1183/13993003.01788-2018 |
| Cluster analysis identifies a pathophysiologically distinct subpopulation with increased serum leptin levels and severe obstructive sleep apnea. | Kozu Y; Kurosawa Y; Yamada S; Fukuda A; Hikichi M; Hiranuma H; Akahoshi T; Gon Y | 2021 | Jun | Sleep Breath | 25 | 2 | 767-776 |  | 10.1007/s11325-020-02160-8 |
| Cluster analysis of clinical phenotypic heterogeneity in obstructive sleep apnea assessed using photoplethysmography. | Zhu W; Xiang L; Long Y; Xun Q; Kuang J; He L | 2023 | Feb | Sleep Med | 102 |  | 134-141 |  | 10.1016/j.sleep.2022.12.023 |
| Cluster Analysis of Home Polygraphic Recordings in Symptomatic Habitually-Snoring Children: A Precision Medicine Perspective. | Zaffanello M; Pietrobelli A; Gozal D; Nosetti L; La Grutta S; Cilluffo G; Ferrante G; Piazza M; Piacentini G | 2022 | Oct | J Clin Med | 11 | 19 |  |  | 10.3390/jcm11195960 |
| Clustering-based characterization of clinical phenotypes in obstructive sleep apnoea using severity, obesity, and craniofacial pattern. | An HJ; Baek SH; Kim SW; Kim SJ; Park YG | 2020 | Jan | Eur J Orthod | 42 | 1 | 93-100 |  | 10.1093/ejo/cjz041 |
| Clusters of sleep apnoea phenotypes: A large pan-European study from the European Sleep Apnoea Database (ESADA). | Bailly S; Grote L; Hedner J; Schiza S; McNicholas WT; Basoglu OK; Lombardi C; Dogas Z; Roisman G; Pataka A; Bonsignore MR; Pepin JL | 2021 | Apr | Respirology | 26 | 4 | 378-387 |  | 10.1111/resp.13969 |
| CMS2-Net: Semi-Supervised Sleep Staging for Diverse Obstructive Sleep Apnea Severity. | Zhang C; Yu W; Li Y; Sun H; Zhang Y; De Vos M | 2022 | Jul | IEEE J Biomed Health Inform | 26 | 7 | 3447-3457 | | 10.1109/JBHI.2022.3156585 |
| Combined unsupervised-supervised machine learning for phenotyping complex diseases with its application to obstructive sleep apnea. | Ma EY; Kim JW; Lee Y; Cho SW; Kim H; Kim JK | 2021 | Feb | Sci Rep | 11 | 1 | 4457 |  | 10.1038/s41598-021-84003-4 |
| Comparative analysis of predictive methods for early assessment of compliance with continuous positive airway pressure therapy. | Rafael-Palou X; Turino C; Steblin A; SÃ¡nchez-de-la-Torre M; BarbÃ© F; Vargiu E | 2018 | Sep | BMC Med Inform Decis Mak | 18 | 1 | 81 | ClinicalTrials.gov/NCT03116958 | 10.1186/s12911-018-0657-z |
| Comparative study of a wearable intelligent sleep monitor and polysomnography monitor for the diagnosis of obstructive sleep apnea. | Xu Y; Ou Q; Cheng Y; Lao M; Pei G | 2023 | Mar | Sleep Breath | 27 | 1 | 205-212 |  | 10.1007/s11325-022-02599-x |
| Comparison of support vector machine based on genetic algorithm with logistic regression to diagnose obstructive sleep apnea. | Manoochehri Z; Salari N; Rezaei M; Khazaie H; Manoochehri S; Pavah BK | 2018 |  | J Res Med Sci | 23 |  | 65 |  | 10.4103/jrms.JRMS_357_17 |
| Comprehensive Analysis of N6-Methyladenosine Regulators in the Subcluster Classification and Drug Candidates Prediction of Severe Obstructive Sleep Apnea. | Li N; Gao Z; Shen J; Liu Y; Wu K; Yang J; Wang S; Zhang X; Zhu Y; Zhu J; Guan J; Liu F; Yin S | 2022 |  | Front Genet | 13 |  | 862972 |  | 10.3389/fgene.2022.862972 |
| Comprehensive Metabolomics and Machine Learning Identify Profound Oxidative Stress and Inflammation Signatures in Hypertensive Patients with Obstructive Sleep Apnea. | Du Z; Sun H; Du Y; Li L; Lv Q; Yu H; Li F; Wang Y; Jiao X; Hu C; Qin Y | 2022 | Sep | Antioxidants (Basel) | 11 | 10 |  |  | 10.3390/antiox11101946 |
| Computational analysis of airflow dynamics for predicting collapsible sites in the upper airways: machine learning approach. | Yeom SH; Na JS; Jung HD; Cho HJ; Choi YJ; Lee JS | 2019 | Oct | J Appl Physiol (1985) | 127 | 4 | 959-973 |  | 10.1152/japplphysiol.01033.2018 |
| Computer Algorithms in Assessment of Obstructive Sleep Apnoea Syndrome and Its Application in Estimating Prevalence of Sleep Related Disorders in Population. | Katyayan A; Yadav V; Mishra P; Mishra A; Saxena M; Kant S; Garg R; Srivastava A; Verma V | 2019 | Sep | Indian J Otolaryngol Head Neck Surg | 71 | 3 | 352-359 |  | 10.1007/s12070-019-01607-z |
| Contactless recording of sleep apnea and periodic leg movements by nocturnal 3-D-video and subsequent visual perceptive computing. | Veauthier C; Ryczewski J; Mansow-Model S; Otte K; Kayser B; Glos M; SchÃ¶bel C; Paul F; Brandt AU; Penzel T | 2019 | Nov | Sci Rep | 9 | 1 | 16812 |  | 10.1038/s41598-019-53050-3 |
| Contribution of Different Subbands of ECG in Sleep Apnea Detection Evaluated Using Filter Bank Decomposition and a Convolutional Neural Network. | Yeh CY; Chang HY; Hu JY; Lin CC | 2022 | Jan | Sensors (Basel) | 22 | 2 |  |  | 10.3390/s22020510 |
| Deep Learning Application to Clinical Decision Support System in Sleep Stage Classification. | Kim D; Lee J; Woo Y; Jeong J; Kim C; Kim DK | 2022 | Jan | J Pers Med | 12 | 2 |  |  | 10.3390/jpm12020136 |
| Deep learning applied to polysomnography to predict blood pressure in obstructive sleep apnea and obesity hypoventilation: a proof-of-concept study. | Prasad B; Agarwal C; Schonfeld E; Schonfeld D; Mokhlesi B | 2020 | Oct | J Clin Sleep Med | 16 | 10 | 1797-1803 | | 10.5664/jcsm.8608 |
| Deep learning enables sleep staging from photoplethysmogram for patients with suspected sleep apnea. | Korkalainen H; Aakko J; Duce B; Kainulainen S; Leino A; Nikkonen S; Afara IO; Myllymaa S; TÃ¶yrÃ¤s J; LeppÃ¤nen T | 2020 | Nov | Sleep | 43 | 11 |  |  | 10.1093/sleep/zsaa098 |
| Deep Learning for Diagnosis and Classification of Obstructive Sleep Apnea: A Nasal Airflow-Based Multi-Resolution Residual Network. | Yue H; Lin Y; Wu Y; Wang Y; Li Y; Guo X; Huang Y; Wen W; Zhao G; Pang X; Lei W | 2021 |  | Nat Sci Sleep | 13 |  | 361-373 |  | 10.2147/NSS.S297856 |
| Deep Learning-Based Assessment of Brain Connectivity Related to Obstructive Sleep Apnea and Daytime Sleepiness. | Lee MH; Lee SK; Thomas RJ; Yoon JE; Yun CH; Shin C | 2021 |  | Nat Sci Sleep | 13 |  | 1561-1572 | | 10.2147/NSS.S327110 |
| Deep-Learning based Sleep Apnea Detection using SpO2 and Pulse Rate. | Sharma P; Jalali A; Majmudar M; Rajput KS; Selvaraj N | 2022 | Jul | Annu Int Conf IEEE Eng Med Biol Soc | 2022 |  | 2611-2614 | | 10.1109/EMBC48229.2022.9871295 |
| Deep-Learning Model Based on Convolutional Neural Networks to Classify Apnea-Hypopnea Events from the Oximetry Signal. | Vaquerizo-Villar F; Ãlvarez D; GutiÃ©rrez-Tobal GC; Arroyo-Domingo CA; Del Campo F; Hornero R | 2022 |  | Adv Exp Med Biol | 1384 |  | 255-264 |  | 10.1007/978-3-031-06413-5_15 |
| Defining the patterns of PAP adherence in pediatric obstructive sleep apnea: a clustering analysis using real-world data. | Weiss MR; Allen ML; Landeo-Gutierrez JS; Lew JP; Aziz JK; Mintz SS; Lawlor CM; Becerra BJ; Preciado DA; Nino G | 2021 | May | J Clin Sleep Med | 17 | 5 | 1005-1013 | | 10.5664/jcsm.9100 |
| Design and Conceptual Proposal of an Intelligent Clinical Decision Support System for the Diagnosis of Suspicious Obstructive Sleep Apnea Patients from Health Profile. | Casal-Guisande M; Torres-DurÃ¡n M; Mosteiro-AÃ±Ã³n M; Cerqueiro-PequeÃ±o J; Bouza-RodrÃ­guez JB; FernÃ¡ndez-Villar A; ComesaÃ±a-Campos A | 2023 | Feb | Int J Environ Res Public Health | 20 | 4 |  |  | 10.3390/ijerph20043627 |
| Design and Evaluation of a Non-ContactBed-Mounted Sensing Device for AutomatedIn-Home Detection of Obstructive Sleep Apnea:A Pilot Study. | Mosquera-Lopez C; Leitschuh J; Condon J; Hagen CC; Rajhbeharrysingh U; Hanks C; Jacobs PG | 2019 | Jul | Biosensors (Basel) | 9 | 3 |  |  | 10.3390/bios9030090 |
| Detailed Assessment of Sleep Architecture With Deep Learning and Shorter Epoch-to-Epoch Duration Reveals Sleep Fragmentation of Patients With Obstructive Sleep Apnea. | Korkalainen H; Leppanen T; Duce B; Kainulainen S; Aakko J; Leino A; Kalevo L; Afara IO; Myllymaa S; Toyras J | 2021 | Jul | IEEE J Biomed Health Inform | 25 | 7 | 2567-2574 | | 10.1109/JBHI.2020.3043507 |
| Detecting inspiratory flow limitation with temporal features of nasal airflow. | Zhi YX; Vena D; Popovic MR; Bradley TD; Yadollahi A | 2018 | Aug | Sleep Med | 48 |  | 70-78 |  | 10.1016/j.sleep.2018.04.006 |
| Detecting obstructive sleep apnea by craniofacial image-based deep learning. | He S; Su H; Li Y; Xu W; Wang X; Han D | 2022 | Dec | Sleep Breath | 26 | 4 | 1885-1895 | | 10.1007/s11325-022-02571-9 |
| Detection of pediatric obstructive sleep apnea using a multilayer perceptron model based on single-channel oxygen saturation or clinical features. | Wu Y; Jia Y; Ning X; Xu Z; Rosen D | 2022 | Aug | Methods | 204 |  | 361-367 |  | 10.1016/j.ymeth.2022.04.017 |
| Detection of Snore from OSAHS Patients Based on Deep Learning. | Shen F; Cheng S; Li Z; Yue K; Li W; Dai L | 2020 |  | J Healthc Eng | 2020 |  | 8864863 |  | 10.1155/2020/8864863 |
| Development and assessment of a risk prediction model for moderate-to-severe obstructive sleep apnea. | Yan X; Wang L; Liang C; Zhang H; Zhao Y; Yu H; Di J | 2022 |  | Front Neurosci | 16 |  | 936946 |  | 10.3389/fnins.2022.936946 |
| Development and validation of moderate to severe obstructive sleep apnea screening test (ColTon) in a pediatric population. | Bokov P; Dudoignon B; Boujemla I; Dahan J; Spruyt K; Delclaux C | 2023 | Apr | Sleep Med | 104 |  | 17-Nov |  | 10.1016/j.sleep.2023.02.016 |
| Development of a Minimally Invasive Screening Tool to Identify Obese Pediatric Population at Risk of Obstructive Sleep Apnea/Hypopnea Syndrome. | CalderÃ³n JM; Ãlvarez-Pitti J; Cuenca I; Ponce F; Redon P | 2020 | Oct | Bioengineering (Basel) | 7 | 4 |  |  | 10.3390/bioengineering7040131 |
| Development of a physiological-based model that uses standard polysomnography and clinical data to predict oral appliance treatment outcomes in obstructive sleep apnea. | Dutta R; Tong BK; Eckert DJ | 2022 | Mar | J Clin Sleep Med | 18 | 3 | 861-870 |  | 10.5664/jcsm.9742 |
| Development of a support vector machine learning and smart phone Internet of Things-based architecture for real-time sleep apnea diagnosis. | Ma B; Wu Z; Li S; Benton R; Li D; Huang Y; Kasukurthi MV; Lin J; Borchert GM; Tan S; Li G; Yang M; Huang J | 2020 | Dec | BMC Med Inform Decis Mak | 20 | Suppl 14 | 298 |  | 10.1186/s12911-020-01329-1 |
| Diagnosis of Obstructive Sleep Apnea during Wakefulness Using Upper Airway Negative Pressure and Machine Learning. | Lim J; Alshaer H; Khan SS; Pandya A; Ryan CM; Haleem A; Sivakulam N; Sahak H; Haq AU; Macarthur K | 2019 | Jul | Annu Int Conf IEEE Eng Med Biol Soc | 2019 |  | 1605-1608 | | 10.1109/EMBC.2019.8856754 |
| Diagnosis of obstructive sleep apnea in children based on the XGBoost algorithm using nocturnal heart rate and blood oxygen feature. | Ye P; Qin H; Zhan X; Wang Z; Liu C; Song B; Kong Y; Jia X; Qi Y; Ji J; Chang L; Ni X; Tai J | 2023 | Mar-Apr | Am J Otolaryngol | 44 | 2 | 103714 |  | 10.1016/j.amjoto.2022.103714 |
| Diagnosis of obstructive sleep apnea with prediction of flow characteristics according to airway morphology automatically extracted from medical images: Computational fluid dynamics and artificial intelligence approach. | Ryu S; Kim JH; Yu H; Jung HD; Chang SW; Park JJ; Hong S; Cho HJ; Choi YJ; Choi J; Lee JS | 2021 | Sep | Comput Methods Programs Biomed | 208 |  | 106243 |  | 10.1016/j.cmpb.2021.106243 |
| Diagnosis of Sleep Apnoea Using a Mandibular Monitor and Machine Learning Analysis: One-Night Agreement Compared to in-Home Polysomnography. | Kelly JL; Ben Messaoud R; Joyeux-Faure M; Terrail R; Tamisier R; Martinot JB; Le-Dong NN; Morrell MJ; PÃ©pin JL | 2022 |  | Front Neurosci | 16 |  | 726880 | ClinicalTrials.gov/NCT04262557 | 10.3389/fnins.2022.726880 |
| Diagnostic Performance of Machine Learning-Derived OSA Prediction Tools in Large Clinical and Community-Based Samples. | Holfinger SJ; Lyons MM; Keenan BT; Mazzotti DR; Mindel J; Maislin G; Cistulli PA; Sutherland K; McArdle N; Singh B; Chen NH; Gislason T; Penzel T; Han F; Li QY; Schwab R; Pack AI; Magalang UJ | 2022 | Mar | Chest | 161 | 3 | 807-817 |  | 10.1016/j.chest.2021.10.023 |
| Distinguishing Obstructive Versus Central Apneas in Infrared Video of Sleep Using Deep Learning: Validation Study. | Akbarian S; Montazeri Ghahjaverestan N; Yadollahi A; Taati B | 2020 | May | J Med Internet Res | 22 | 5 | e17252 |  | 10.2196/17252 |
| Dreem Open Datasets: Multi-Scored Sleep Datasets to Compare Human and Automated Sleep Staging. | Guillot A; Sauvet F; During EH; Thorey V | 2020 | Sep | IEEE Trans Neural Syst Rehabil Eng | 28 | 9 | 1955-1965 | | 10.1109/TNSRE.2020.3011181 |
| Empirical Analysis of Apnea Syndrome Using an Artificial Intelligence-Based Granger Panel Model Approach. | Onyema EM; Ahanger TA; Samir G; Shrivastava M; Maheshwari M; Seghir GM; Krah D | 2022 |  | Comput Intell Neurosci | 2022 |  | 7969389 |  | 10.1155/2022/7969389 |
| Endotyping Sleep Apnea One Breath at a Time: An Automated Approach for Separating Obstructive from Central Sleep-disordered Breathing. | Parekh A; Tolbert TM; Mooney AM; Ramos-Cejudo J; Osorio RS; Treml M; Herkenrath SD; Randerath WJ; Ayappa I; Rapoport DM | 2021 | Dec | Am J Respir Crit Care Med | 204 | 12 | 1452-1462 | | 10.1164/rccm.202011-4055OC |
| Enhancing Obstructive Sleep Apnea Diagnosis With Screening Through Disease Phenotypes: Algorithm Development and Validation. | Ferreira-Santos D; Rodrigues PP | 2021 | Jun | JMIR Med Inform | 9 | 6 | e25124 |  | 10.2196/25124 |
| Ensemble of Deep Learning Models for Sleep Apnea Detection: An Experimental Study. | Mukherjee D; Dhar K; Schwenker F; Sarkar R | 2021 | Aug | Sensors (Basel) | 21 | 16 |  |  | 10.3390/s21165425 |
| Estimating daytime sleepiness with previous night electroencephalography, electrooculography, and electromyography spectrograms in patients with suspected sleep apnea using a convolutional neural network. | Nikkonen S; Korkalainen H; Kainulainen S; Myllymaa S; Leino A; Kalevo L; Oksenberg A; LeppÃ¤nen T; TÃ¶yrÃ¤s J | 2020 | Dec | Sleep | 43 | 12 |  |  | 10.1093/sleep/zsaa106 |
| Estimation of Apnea-Hypopnea Index Using Deep Learning On 3-D Craniofacial Scans. | Hanif U; Leary E; Schneider L; Paulsen R; Morse AM; Blackman A; Schweitzer P; Kushida CA; Liu S; Jennum P; Sorensen H; Mignot E | 2021 | Nov | IEEE J Biomed Health Inform | 25 | 11 | 4185-4194 | | 10.1109/JBHI.2021.3078127 |
| Estimation of cerebral blood flow velocity during breath-hold challenge using artificial neural networks. | Al-Abed MA; Al-Bashir AK; Al-Rawashdeh A; Alex RM; Zhang R; Watenpaugh DE; Behbehani K | 2019 | Dec | Comput Biol Med | 115 |  | 103508 |  | 10.1016/j.compbiomed.2019.103508 |
| Evaluating an under-mattress sleep monitor compared to a peripheral arterial tonometry home sleep apnea test device in the diagnosis of obstructive sleep apnea. | Jagielski JT; Bibi N; Gay PC; Junna MR; Carvalho DZ; Williams JA; Morgenthaler TI | 2022 | Nov | Sleep Breath | |  |  | ClinicalTrials.gov/NCT04778748 | 10.1007/s11325-022-02751-7 |
| Evaluating Prediction Models of Sleep Apnea From Smartphone-Recorded Sleep Breathing Sounds. | Cho SW; Jung SJ; Shin JH; Won TB; Rhee CS; Kim JW | 2022 | Jun | JAMA Otolaryngol Head Neck Surg | 148 | 6 | 515-521 |  | 10.1001/jamaoto.2022.0244 |
| Evaluation of obstructive sleep apnea: an analysis based on aberrant genes. | Liao J; Gao X; Shi Y; Li Y; Han D | 2022 | Nov | Sleep Breath | |  |  |  | 10.1007/s11325-022-02749-1 |
| Explainable fuzzy neural network with easy-to-obtain physiological features for screening obstructive sleep apnea-hypopnea syndrome. | Juang CF; Wen CY; Chang KM; Chen YH; Wu MF; Huang WC | 2021 | Sep | Sleep Med | 85 |  | 280-290 |  | 10.1016/j.sleep.2021.07.012 |
| Feasibility of Single Channel Oximetry for Mass Screening of Obstructive Sleep Apnea. | Behar JA; Palmius N; Li Q; Garbuio S; Rizzatti FPG; Bittencourt L; Tufik S; Clifford GD | 2019 | May-Jun | EClinicalMedicine | 11 |  | 81-88 |  | 10.1016/j.eclinm.2019.05.015 |
| Feature relevance in physiological networks for classification of obstructive sleep apnea. | Jansen C; Hodel S; Penzel T; Spott M; Krefting D | 2018 | Dec | Physiol Meas | 39 | 12 | 124003 |  | 10.1088/1361-6579/aaf0c9 |
| Frequency Network Analysis of Heart Rate Variability for Obstructive Apnea Patient Detection. | Dong Z; Li X; Chen W | 2018 | Nov | IEEE J Biomed Health Inform | 22 | 6 | 1895-1905 | | 10.1109/JBHI.2017.2784415 |
| Gender Phenotyping of Patients with Obstructive Sleep Apnea Syndrome Using a Network Science Approach. | TopÃ®rceanu A; Udrescu L; Udrescu M; Mihaicuta S | 2020 | Dec | J Clin Med | 9 | 12 |  |  | 10.3390/jcm9124025 |
| Hidden Markov model segmentation to demarcate trajectories of residual apnoea-hypopnoea index in CPAP-treated sleep apnoea patients to personalize follow-up and prevent treatment failure. | Midelet A; Bailly S; Tamisier R; Borel JC; Baillieul S; Le Hy R; Schaeffer MC; PÃ©pin JL | 2021 | Dec | EPMA J | 12 | 4 | 535-544 |  | 10.1007/s13167-021-00264-z |
| Homecare interventions as a Service model for Obstructive sleep Apnea: Delivering personalised phone call using patient profiling and adherence predictions. | Joymangul JS; Sekhari A; Grasset O; Moalla N | 2023 | Feb | Int J Med Inform | 170 |  | 104935 |  | 10.1016/j.ijmedinf.2022.104935 |
| Identification and Validation of Prognostic Factors of Lipid Metabolism in Obstructive Sleep Apnea. | Peng L; Wang X; Bing D | 2021 |  | Front Genet | 12 |  | 747576 |  | 10.3389/fgene.2021.747576 |
| Identification of key genes and immune infiltration modulated by CPAP in obstructive sleep apnea by integrated bioinformatics analysis. | Fan C; Huang S; Xiang C; An T; Song Y | 2021 |  | PLoS One | 16 | 9 | e0255708 |  | 10.1371/journal.pone.0255708 |
| Identification of the Novel Gene Markers Based on the Gene Profile among Different Severity of Obstructive Sleep Apnea. | Ren Y; Li Y; Sui X; Yuan J; Lan J; Li X; Deng Y; Xu Z; Cheng X; Zhao C; Lu J | 2022 |  | Comput Math Methods Med | 2022 |  | 6517965 |  | 10.1155/2022/6517965 |
| In obstructive sleep apnea patients, automatic determination of respiratory arrests by photoplethysmography signal and heart rate variability. | Bozkurt MR; UÃ§ar MK; Bozkurt F; Bilgin C | 2019 | Dec | Australas Phys Eng Sci Med | 42 | 4 | 959-979 |  | 10.1007/s13246-019-00796-9 |
| Inherent regional brain activity changes in male obstructive sleep apnea with mild cognitive impairment: A resting-state magnetic resonance study. | Shu Y; Liu X; Yu P; Li H; Duan W; Wei Z; Li K; Xie W; Zeng Y; Peng D | 2022 |  | Front Aging Neurosci | 14 |  | 1022628 |  | 10.3389/fnagi.2022.1022628 |
| In-home mandibular repositioning during sleep using MATRx plus predicts outcome and efficacious positioning for oral appliance treatment of obstructive sleep apnea. | Mosca EV; Bruehlmann S; Zouboules SM; Chiew AE; Westersund C; Hambrook DA; Jahromi SAZ; Grosse J; Topor ZL; Charkhandeh S; Remmers JE | 2022 | Mar | J Clin Sleep Med | 18 | 3 | 911-919 | ClinicalTrials.gov/NCT03217383 | 10.5664/jcsm.9758 |
| Integrating domain knowledge with machine learning to detect obstructive sleep apnea: Snore as a significant bio-feature. | Hsu YC; Wang JD; Huang PH; Chien YW; Chiu CJ; Lin CY | 2022 | Apr | J Sleep Res | 31 | 2 | e13487 |  | 10.1111/jsr.13487 |
| Integrating the STOP-BANG Score and Clinical Data to Predict Cardiovascular Events After Infarction: A Machine Learning Study. | Calvillo-ArgÃ¼elles O; Sierra-FernÃ¡ndez CR; Padilla-Ibarra J; Rodriguez-Zanella H; Balderas-MuÃ±oz K; Arias-Mendoza MA; MartÃ­nez-SÃ¡nchez C; Selmen-Chattaj S; Dominguez-Mendez BE; van der Harst P; Juarez-Orozco LE | 2020 | Oct | Chest | 158 | 4 | 1669-1679 | | 10.1016/j.chest.2020.03.074 |
| Introducing the Hybrid "K-means, RLS" Learning for the RBF Network in Obstructive Apnea Disease Detection using Dual-tree Complex Wavelet Transform Based Features. | Ostadieh J; Amirani MC | 2020 | Jan | J Electr Bioimpedance | 11 | 1 | 11-Apr |  | 10.2478/joeb-2020-0002 |
| Investigation on factors related to poor CPAP adherence using machine learning: a pilot study. | Eguchi K; Yabuuchi T; Nambu M; Takeyama H; Azuma S; Chin K; Kuroda T | 2022 | Nov | Sci Rep | 12 | 1 | 19563 |  | 10.1038/s41598-022-21932-8 |
| Logistic regression and artificial neural network-based simple predicting models for obstructive sleep apnea by age, sex, and body mass index. | Kuan YC; Hong CT; Chen PC; Liu WT; Chung CC | 2022 | Aug | Math Biosci Eng | 19 | 11 | 11409-11421 | | 10.3934/mbe.2022532 |
| Low Level Texture Features for Snore Sound Discrimination. | Demir F; Sengur A; Cummins N; Amiriparian S; Schuller B | 2018 | Jul | Annu Int Conf IEEE Eng Med Biol Soc | 2018 |  | 413-416 |  | 10.1109/EMBC.2018.8512459 |
| Machine learning and geometric morphometrics to predict obstructive sleep apnea from 3D craniofacial scans. | Monna F; Ben Messaoud R; Navarro N; Baillieul S; Sanchez L; Loiodice C; Tamisier R; Joyeux-Faure M; PÃ©pin JL | 2022 | Jul | Sleep Med | 95 |  | 76-83 | ClinicalTrials.gov/NCT03632382 | 10.1016/j.sleep.2022.04.019 |
| Machine learning approach for obstructive sleep apnea screening using brain diffusion tensor imaging. | Pang B; Doshi S; Roy B; Lai M; Ehlert L; Aysola RS; Kang DW; Anderson A; Joshi SH; Tward D; Scalzo F; Vacas S; Kumar R | 2023 | Feb | J Sleep Res | 32 | 1 | e13729 |  | 10.1111/jsr.13729 |
| Machine learning approaches for screening the risk of obstructive sleep apnea in the Taiwan population based on body profile. | Tsai CY; Liu WT; Lin YT; Lin SY; Houghton R; Hsu WH; Wu D; Lee HC; Wu CJ; Li LYJ; Hsu SM; Lo CC; Lo K; Chen YR; Lin FC; Majumdar A | 2022 | Oct | Inform Health Soc Care | 47 | 4 | 373-388 |  | 10.1080/17538157.2021.2007930 |
| Machine learning for image-based detection of patients with obstructive sleep apnea: an exploratory study. | Tsuiki S; Nagaoka T; Fukuda T; Sakamoto Y; Almeida FR; Nakayama H; Inoue Y; Enno H | 2021 | Dec | Sleep Breath | 25 | 4 | 2297-2305 | | 10.1007/s11325-021-02301-7 |
| Machine learning for nocturnal mass diagnosis of atrial fibrillation in a population at risk of sleep-disordered breathing. | Chocron A; Efraim R; Mandel F; Rueschman M; Palmius N; Penzel T; Elbaz M; Behar JA | 2020 | Nov | Physiol Meas | 41 | 10 | 104001 |  | 10.1088/1361-6579/abb8bf |
| Machine Learning Identification of Obstructive Sleep Apnea Severity through the Patient Clinical Features: A Retrospective Study. | Maniaci A; Riela PM; Iannella G; Lechien JR; La Mantia I; De Vincentiis M; Cammaroto G; Calvo-Henriquez C; Di Luca M; Chiesa Estomba C; Saibene AM; Pollicina I; Stilo G; Di Mauro P; Cannavicci A; Lugo R; Magliulo G; Greco A; Pace A; Meccariello G; Cocuzza S; Vicini C | 2023 | Mar | Life (Basel) | 13 | 3 |  |  | 10.3390/life13030702 |
| Machine learning-based model for prediction of outcomes in palatal surgery for obstructive sleep apnoea. | Yang SJ; Kim JS; Chung SK; Song YY | 2021 | Nov | Clin Otolaryngol | 46 | 6 | 1242-1246 | | 10.1111/coa.13823 |
| Machine learning-based prediction of adherence to continuous positive airway pressure (CPAP) in obstructive sleep apnea (OSA). | Scioscia G; Tondo P; Foschino Barbaro MP; Sabato R; Gallo C; Maci F; Lacedonia D | 2022 | Jul | Inform Health Soc Care | 47 | 3 | 274-282 |  | 10.1080/17538157.2021.1990300 |
| Machine learning-based preoperative datamining can predict the therapeutic outcome of sleep surgery in OSA subjects. | Kim JY; Kong HJ; Kim SH; Lee S; Kang SH; Han SC; Kim DW; Ji JY; Kim HJ | 2021 | Jul | Sci Rep | 11 | 1 | 14911 |  | 10.1038/s41598-021-94454-4 |
| Management and Treatment of Patients With Obstructive Sleep Apnea Using an Intelligent Monitoring System Based on Machine Learning Aiming to Improve Continuous Positive Airway Pressure Treatment Compliance: Randomized Controlled Trial. | Turino C; BenÃ­tez ID; Rafael-Palou X; Mayoral A; Lopera A; Pascual L; Vaca R; Cortijo A; MoncusÃ­-Moix A; Dalmases M; Vargiu E; Blanco J; BarbÃ© F; de Batlle J | 2021 | Oct | J Med Internet Res | 23 | 10 | e24072 | ClinicalTrials.gov/NCT03116958 | 10.2196/24072 |
| Moderate to severe OSA screening based on support vector machine of the Chinese population faciocervical measurements dataset: a cross-sectional study. | Zhang L; Yan YR; Li SQ; Li HP; Lin YN; Li N; Sun XW; Ding YJ; Li CX; Li QY | 2021 | Sep | BMJ Open | 11 | 9 | e048482 |  | 10.1136/bmjopen-2020-048482 |
| Mortality Patterns Associated with Central Sleep Apnea among Veterans: A Large, Retrospective, Longitudinal Report. | Agrawal R; Sharafkhaneh A; Gottlieb DJ; Nowakowski S; Razjouyan J | 2023 | Mar | Ann Am Thorac Soc | 20 | 3 | 450-455 |  | 10.1513/AnnalsATS.202207-648OC |
| Multiclass classification of obstructive sleep apnea/hypopnea based on a convolutional neural network from a single-lead electrocardiogram. | Urtnasan E; Park JU; Lee KJ | 2018 | Jun | Physiol Meas | 39 | 6 | 65003 |  | 10.1088/1361-6579/aac7b7 |
| Multi-perspective clustering of obstructive sleep apnea towards precision therapeutic decision including craniofacial intervention. | Kim SJ; Alnakhli WM; Alfaraj AS; Kim KA; Kim SW; Liu SY | 2021 | Mar | Sleep Breath | 25 | 1 | 85-94 |  | 10.1007/s11325-020-02062-9 |
| Multiple Machine Learning Methods Reveal Key Biomarkers of Obstructive Sleep Apnea and Continuous Positive Airway Pressure Treatment. | Zhu J; Sanford LD; Ren R; Zhang Y; Tang X | 2022 |  | Front Genet | 13 |  | 927545 |  | 10.3389/fgene.2022.927545 |
| Neural network analysis of nocturnal SpO(2) signal enables easy screening of sleep apnea in patients with acute cerebrovascular disease. | Leino A; Nikkonen S; Kainulainen S; Korkalainen H; TÃ¶yrÃ¤s J; Myllymaa S; LeppÃ¤nen T; YlÃ¤-Herttuala S; Westeren-Punnonen S; Muraja-Murro A; JÃ¤kÃ¤lÃ¤ P; Mervaala E; Myllymaa K | 2021 | Mar | Sleep Med | 79 |  | 71-78 |  | 10.1016/j.sleep.2020.12.032 |
| Noncontact Sleep Monitoring With Infrared Video Data to Estimate Sleep Apnea Severity and Distinguish Between Positional and Nonpositional Sleep Apnea: Model Development and Experimental Validation. | Akbarian S; Ghahjaverestan NM; Yadollahi A; Taati B | 2021 | Nov | J Med Internet Res | 23 | 11 | e26524 |  | 10.2196/26524 |
| Non-invasive machine learning estimation of effort differentiates sleep-disordered breathing pathology. | Hanif U; Schneider LD; Trap L; Leary EB; Moore H; Guilleminault C; Jennum P; Sorensen HBD; Mignot EJM | 2019 | Feb | Physiol Meas | 40 | 2 | 25008 |  | 10.1088/1361-6579/ab0559 |
| Objective Pharyngeal Phenotyping in Obstructive Sleep Apnea With High-Resolution Manometry. | Kent DT; Scott WC; Ye C; Fabbri D | 2023 | Jan | Otolaryngol Head Neck Surg | | |  |  | 10.1002/ohn.257 |
| Objective Relationship Between Sleep Apnea and Frequency of Snoring Assessed by Machine Learning. | Alshaer H; Hummel R; Mendelson M; Marshal T; Bradley TD | 2019 | Mar | J Clin Sleep Med | 15 | 3 | 463-470 |  | 10.5664/jcsm.7676 |
| Obstructive Sleep Apnea Detection Based on Sleep Sounds via Deep Learning. | Wang B; Tang X; Ai H; Li Y; Xu W; Wang X; Han D | 2022 |  | Nat Sci Sleep | 14 |  | 2033-2045 | | 10.2147/NSS.S373367 |
| Obstructive sleep apnea detection from single-lead electrocardiogram signals using one-dimensional squeeze-and-excitation residual group network. | Yang Q; Zou L; Wei K; Liu G | 2021 | Dec | Comput Biol Med | 140 |  | 105124 |  | 10.1016/j.compbiomed.2021.105124 |
| Obstructive Sleep Apnea Detection Scheme Based on Manually Generated Features andParallel Heterogeneous Deep Learning Model under IoMT. | Shao S; Han G; Wang T; Song C; Yao C; Hou J | 2022 | Apr | IEEE J Biomed Health Inform | PP |  |  |  | 10.1109/JBHI.2022.3166859 |
| Obstructive sleep apnea detection using discrete wavelet transform-based statistical features. | Rajesh KNVPS; Dhuli R; Kumar TS | 2021 | Mar | Comput Biol Med | 130 |  | 104199 |  | 10.1016/j.compbiomed.2020.104199 |
| Obstructive Sleep Apnea Detection using Frequency Analysis of Electrocardiographic RR Interval and Machine Learning Algorithms. | Indrawati AN; Nuryani N; Nugroho AS; Utomo TP | 2022 | Dec | J Biomed Phys Eng | 12 | 6 | 627-636 |  | 10.31661/jbpe.v0i0.2010-1216 |
| Obstructive sleep apnea phenotypes in men based on characteristics of respiratory events during polysomnography. | Nakayama H; Kobayashi M; Tsuiki S; Yanagihara M; Inoue Y | 2019 | Dec | Sleep Breath | 23 | 4 | 1087-1094 | | 10.1007/s11325-019-01785-8 |
| Obstructive sleep apnea prediction from electrocardiogram scalograms and spectrograms using convolutional neural networks. | Nasifoglu H; Erogul O | 2021 | Jun | Physiol Meas | 42 | 6 |  |  | 10.1088/1361-6579/ac0a9c |
| Obstructive sleep apnea predicts 10-year cardiovascular disease-related mortality in the Sleep Heart Health Study: a machine learning approach. | Li A; Roveda JM; Powers LS; Quan SF | 2022 | Feb | J Clin Sleep Med | 18 | 2 | 497-504 |  | 10.5664/jcsm.9630 |
| Obstructive Sleep Apnea Recognition Based on Multi-Bands Spectral Entropy Analysis of Short-Time Heart Rate Variability. | Shao S; Wang T; Song C; Chen X; Cui E; Zhao H | 2019 | Aug | Entropy (Basel) | 21 | 8 |  |  | 10.3390/e21080812 |
| Obstructive sleep apnea screening by heart rate variability-based apnea/normal respiration discriminant model. | Nakayama C; Fujiwara K; Sumi Y; Matsuo M; Kano M; Kadotani H | 2019 | Dec | Physiol Meas | 40 | 12 | 125001 |  | 10.1088/1361-6579/ab57be |
| Obstructive sleep apnea syndrome detection based on ballistocardiogram via machine learning approach. | Gao WD; Xu YB; Li SS; Fu YJ; Zheng DY; She YJ | 2019 | Jun | Math Biosci Eng | 16 | 5 | 5672-5686 | | 10.3934/mbe.2019282 |
| Obstructive Sleep Apnea Syndrome Treated Using a Positive Pressure Ventilator Based on Artificial Intelligence Processor. | Chen Z; Zhao Z; Zhang Z | 2021 |  | J Healthc Eng | 2021 |  | 5683433 |  | 10.1155/2021/5683433 |
| Obstructive sleep apnea: A categorical cluster analysis and visualization. | Ferreira-Santos D; Rodrigues PP | 2021 | Nov | Pulmonology | |  |  |  | 10.1016/j.pulmoe.2021.10.003 |
| Obstructive Sleep Apnea: A Prediction Model Using Supervised Machine Learning Method. | Keshavarz Z; Rezaee R; Nasiri M; Pournik O | 2020 | Jun | Stud Health Technol Inform | 272 |  | 387-390 |  | 10.3233/SHTI200576 |
| Obstructive Sleep Apnoea Syndrome Screening Through Wrist-Worn Smartbands: A Machine-Learning Approach. | Benedetti D; Olcese U; Bruno S; Barsotti M; Maestri Tassoni M; Bonanni E; Siciliano G; Faraguna U | 2022 |  | Nat Sci Sleep | 14 |  | 941-956 |  | 10.2147/NSS.S352335 |
| Orthogonal convolutional neural networks for automatic sleep stage classification based on single-channel EEG. | Zhang J; Yao R; Ge W; Gao J | 2020 | Jan | Comput Methods Programs Biomed | 183 |  | 105089 |  | 10.1016/j.cmpb.2019.105089 |
| Overnight airway obstruction severity prediction centered on acoustic properties of smart phone: validation with esophageal pressure. | Markandeya MN; Abeyratne UR; Hukins C | 2020 | Nov | Physiol Meas | 41 | 10 | 105002 |  | 10.1088/1361-6579/abb75f |
| Paediatric sleep apnea event prediction using nasal air pressure and machine learning. | Crowson MG; Gipson KS; Kadosh OK; Hartnick E; Grealish E; Keamy DG; Kinane TB; Hartnick CJ | 2023 | Feb | J Sleep Res | |  | e13851 |  | 10.1111/jsr.13851 |
| Pediatric Automatic Sleep Staging: A Comparative Study of State-of-the-Art Deep Learning Methods. | Phan H; Mertins A; Baumert M | 2022 | Dec | IEEE Trans Biomed Eng | 69 | 12 | 3612-3622 | | 10.1109/TBME.2022.3174680 |
| Pediatric sleep apnea: Characterization of apneic events and sleep stages using heart rate variability. | MartÃ­n-Montero A; ArmaÃ±ac-JuliÃ¡n P; Gil E; Kheirandish-Gozal L; Ãlvarez D; LÃ¡zaro J; BailÃ³n R; Gozal D; Laguna P; Hornero R; GutiÃ©rrez-Tobal GC | 2023 | Mar | Comput Biol Med | 154 |  | 106549 |  | 10.1016/j.compbiomed.2023.106549 |
| Performance evaluation of the spectral autocorrelation function and autoregressive models for automated sleep apnea detection using single-lead ECG signal. | Zarei A; Mohammadzadeh Asl B | 2020 | Oct | Comput Methods Programs Biomed | 195 |  | 105626 |  | 10.1016/j.cmpb.2020.105626 |
| Performance of facial expression classification tasks in patients with obstructive sleep apnea. | Guo J; Ma Y; Liu Z; Wang F; Hou X; Chen J; Hong Y; Xu S; Liu X | 2020 | Apr | J Clin Sleep Med | 16 | 4 | 523-530 |  | 10.5664/jcsm.8256 |
| Plasma profiling reveals a blood-based metabolic fingerprint of obstructive sleep apnea. | Pinilla L; BenÃ­tez ID; Santamaria-Martos F; Targa A; MoncusÃ­-Moix A; Dalmases M; MÃ­nguez O; AguilÃ  M; JovÃ© M; Sol J; Pamplona R; BarbÃ© F; SÃ¡nchez-de-la-Torre M | 2022 | Jan | Biomed Pharmacother | 145 |  | 112425 |  | 10.1016/j.biopha.2021.112425 |
| Point-of-care prediction model of loop gain in patients with obstructive sleep apnea: development and validation. | Schmickl CN; Orr JE; Kim P; Nokes B; Sands S; Manoharan S; McGinnis L; Parra G; DeYoung P; Owens RL; Malhotra A | 2022 | Apr | BMC Pulm Med | 22 | 1 | 158 |  | 10.1186/s12890-022-01950-y |
| Polysomnographic phenotypes and their cardiovascular implications in obstructive sleep apnoea. | Zinchuk AV; Jeon S; Koo BB; Yan X; Bravata DM; Qin L; Selim BJ; Strohl KP; Redeker NS; Concato J; Yaggi HK | 2018 | May | Thorax | 73 | 5 | 472-480 |  | 10.1136/thoraxjnl-2017-210431 |
| Polysomnographic phenotyping of obstructive sleep apnea and its implications in mortality in Korea. | Kim JW; Won TB; Rhee CS; Park YM; Yoon IY; Cho SW | 2020 | Aug | Sci Rep | 10 | 1 | 13207 |  | 10.1038/s41598-020-70039-5 |
| Portable Sleep Apnea Syndrome Screening and Event Detection Using Long Short-Term Memory Recurrent Neural Network. | Chang HC; Wu HT; Huang PC; Ma HP; Lo YL; Huang YH | 2020 | Oct | Sensors (Basel) | 20 | 21 |  |  | 10.3390/s20216067 |
| Power spectral densities of nocturnal pulse oximetry signals differ in OSA patients with and without daytime sleepiness. | Kainulainen S; TÃ¶yrÃ¤s J; Oksenberg A; Korkalainen H; Afara IO; Leino A; Kalevo L; Nikkonen S; Gadoth N; Kulkas A; Myllymaa S; LeppÃ¤nen T | 2020 | Sep | Sleep Med | 73 |  | 231-237 |  | 10.1016/j.sleep.2020.07.015 |
| Predicting Nondiagnostic Home Sleep Apnea Tests Using Machine Learning. | Stretch R; Ryden A; Fung CH; Martires J; Liu S; Balasubramanian V; Saedi B; Hwang D; Martin JL; Della Penna N; Zeidler MR | 2019 | Nov | J Clin Sleep Med | 15 | 11 | 1599-1608 | | 10.5664/jcsm.8020 |
| Predicting polysomnographic severity thresholds in children using machine learning. | Bertoni D; Sterni LM; Pereira KD; Das G; Isaiah A | 2020 | Sep | Pediatr Res | 88 | 3 | 404-411 |  | 10.1038/s41390-020-0944-0 |
| Predicting Polysomnography Parameters from Anthropometric Features and Breathing Sounds Recorded during Wakefulness. | Elwali A; Moussavi Z | 2021 | May | Diagnostics (Basel) | 11 | 5 |  |  | 10.3390/diagnostics11050905 |
| Prediction model of obstructive sleep apnea-related hypertension: Machine learning-based development and interpretation study. | Shi Y; Ma L; Chen X; Li W; Feng Y; Zhang Y; Cao Z; Yuan Y; Xie Y; Liu H; Yin L; Zhao C; Wu S; Ren X | 2022 |  | Front Cardiovasc Med | 9 |  | 1042996 |  | 10.3389/fcvm.2022.1042996 |
| Prediction of Apnea-Hypopnea Index Using Sound Data Collected by a Noncontact Device. | Kim JW; Kim T; Shin J; Lee K; Choi S; Cho SW | 2020 | Mar | Otolaryngol Head Neck Surg | 162 | 3 | 392-399 |  | 10.1177/0194599819900014 |
| Prediction of Obstructive Sleep Apnea Based on Respiratory Sounds Recorded Between Sleep Onset and Sleep Offset. | Kim JW; Kim T; Shin J; Choe G; Lim HJ; Rhee CS; Lee K; Cho SW | 2019 | Feb | Clin Exp Otorhinolaryngol | 12 | 1 | 72-78 |  | 10.21053/ceo.2018.00388 |
| Prediction of obstructive sleep apnea using deep learning in 3D craniofacial reconstruction. | Zhang Z; Feng Y; Li Y; Zhao L; Wang X; Han D | 2023 | Jan | J Thorac Dis | 15 | 1 | 90-100 |  | 10.21037/jtd-22-734 |
| Prediction of obstructive sleep apnea using ensemble of recurrence plot convolutional neural networks (RPCNNs) from polysomnography signals. | Taghizadegan Y; Jafarnia Dabanloo N; Maghooli K; Sheikhani A | 2021 | Sep | Med Hypotheses | 154 |  | 110659 |  | 10.1016/j.mehy.2021.110659 |
| Prediction of Oxygen Desaturation by Using Sound Data From a Noncontact Device: A Proof-of-Concept Study. | Kim JW; Shin J; Lee K; Won TB; Rhee CS; Cho SW | 2022 | Apr | Laryngoscope | 132 | 4 | 901-905 |  | 10.1002/lary.29971 |
| Predictive performances of 6 data mining techniques for obstructive sleep apnea-hypopnea syndrome. | Luo M; Feng Y; Luo J; Li X; Han J; Li T | 2022 | Jul | Medicine (Baltimore) | 101 | 26 | e29724 |  | 10.1097/MD.0000000000029724 |
| Probabilistic domain-knowledge modeling of disorder pathogenesis for dynamics forecasting of acute onset. | Huynh PK; Setty A; Phan H; Le TQ | 2021 | May | Artif Intell Med | 115 |  | 102056 |  | 10.1016/j.artmed.2021.102056 |
| Proteomic biomarkers of sleep apnea. | Ambati A; Ju YE; Lin L; Olesen AN; Koch H; Hedou JJ; Leary EB; Sempere VP; Mignot E; Taheri S | 2020 | Nov | Sleep | 43 | 11 |  |  | 10.1093/sleep/zsaa086 |
| Proteomic Biomarkers of the Apnea Hypopnea Index and Obstructive Sleep Apnea: Insights into the Pathophysiology of Presence, Severity, and Treatment Response. | Cederberg KLJ; Hanif U; Peris Sempere V; HÃ©dou J; Leary EB; Schneider LD; Lin L; Zhang J; Morse AM; Blackman A; Schweitzer PK; Kotagal S; Bogan R; Kushida CA; Ju YS; Petousi N; Turnbull CD; Mignot E; The Stages Cohort Investigator Group | 2022 | Jul | Int J Mol Sci | 23 | 14 |  |  | 10.3390/ijms23147983 |
| Proteomic profiling for prediction of recurrent cardiovascular event in patients with acute coronary syndrome and obstructive sleep apnea: A post-hoc analysis from the ISAACC study. | Zapater A; Gracia-Lavedan E; Torres G; MÃ­nguez O; Pascual L; Cortijo A; MartÃ­nez D; BenÃ­tez ID; De Batlle J; HenrÃ­quez-BeltrÃ¡n M; Abad J; Duran-Cantolla J; Urrutia A; Mediano O; Masdeu MJ; Ordax-Carbajo E; Masa JF; De la PeÃ±a M; Mayos M; Coloma R; Montserrat JM; Chiner E; BarbÃ© F; SÃ¡nchez-de-la-Torre M | 2023 | Feb | Biomed Pharmacother | 158 |  | 114125 |  | 10.1016/j.biopha.2022.114125 |
| Radar-based sleep stage classification in children undergoing polysomnography: a pilot-study. | de Goederen R; Pu S; Silos Viu M; Doan D; Overeem S; Serdijn WA; Joosten KFM; Long X; Dudink J | 2021 | Jun | Sleep Med | 82 |  | 8-Jan |  | 10.1016/j.sleep.2021.03.022 |
| Real-Time Detection of Sleep Apnea Based on Breathing Sounds and Prediction Reinforcement Using Home Noises: Algorithm Development and Validation. | Le VL; Kim D; Cho E; Jang H; Reyes RD; Kim H; Lee D; Yoon IY; Hong J; Kim JW | 2023 | Feb | J Med Internet Res | 25 |  | e44818 |  | 10.2196/44818 |
| Real-time prediction of upcoming respiratory events via machine learning using snoring sound signal. | Wang B; Yi X; Gao J; Li Y; Xu W; Wu J; Han D | 2021 | Sep | J Clin Sleep Med | 17 | 9 | 1777-1784 | | 10.5664/jcsm.9292 |
| Recognition of Patient Groups with Sleep Related Disorders using Bio-signal Processing and Deep Learning. | Jarchi D; Andreu-Perez J; Kiani M; Vysata O; Kuchynka J; Prochazka A; Sanei S | 2020 | May | Sensors (Basel) | 20 | 9 |  |  | 10.3390/s20092594 |
| Regional characterization of functional connectivity in patients with sleep apnea hypopnea syndrome during sleep. | Zhang T; Pan Y; Lian J; Pang F; Wen J; Luo Y | 2021 | Jul | Physiol Meas | 42 | 7 |  |  | 10.1088/1361-6579/ac0e83 |
| Respiratory effort during sleep and prevalent hypertension in obstructive sleep apnoea. | Martinot JB; Le-Dong NN; Malhotra A; PÃ©pin JL | 2023 | Mar | Eur Respir J | 61 | 3 |  |  | 10.1183/13993003.01486-2022 |
| SCNN: Scalogram-based convolutional neural network to detect obstructive sleep apnea using single-lead electrocardiogram signals. | Mashrur FR; Islam MS; Saha DK; Islam SMR; Moni MA | 2021 | Jul | Comput Biol Med | 134 |  | 104532 |  | 10.1016/j.compbiomed.2021.104532 |
| Screening for Obstructive Sleep Apnea Risk by Using Machine Learning Approaches and Anthropometric Features. | Tsai CY; Huang HT; Cheng HC; Wang J; Duh PJ; Hsu WH; Stettler M; Kuan YC; Lin YT; Hsu CR; Lee KY; Kang JH; Wu D; Lee HC; Wu CJ; Majumdar A; Liu WT | 2022 | Nov | Sensors (Basel) | 22 | 22 |  |  | 10.3390/s22228630 |
| Screening for obstructive sleep apnea with novel hybrid acoustic smartphone app technology. | Tiron R; Lyon G; Kilroy H; Osman A; Kelly N; O'Mahony N; Lopes C; Coffey S; McMahon S; Wren M; Conway K; Fox N; Costello J; Shouldice R; Lederer K; Fietze I; Penzel T | 2020 | Aug | J Thorac Dis | 12 | 8 | 4476-4495 | | 10.21037/jtd-20-804 |
| Screening the risk of obstructive sleep apnea by utilizing supervised learning techniques based on anthropometric features and snoring events. | Tsai CY; Liu WT; Hsu WH; Majumdar A; Stettler M; Lee KY; Cheng WH; Wu D; Lee HC; Kuan YC; Wu CJ; Lin YC; Ho SC | 2023 | Jan-Dec | Digit Health | 9 |  | 2.06E+16 |  | 10.1177/20552076231152751 |
| Selection of OSA-specific pronunciations and assessment of disease severity assisted by machine learning. | Ding Y; Sun Y; Li Y; Wang H; Fang Q; Xu W; Wu J; Gao J; Han D | 2022 | Nov | J Clin Sleep Med | 18 | 11 | 2663-2672 | | 10.5664/jcsm.9798 |
| Severity evaluation of obstructive sleep apnea based on speech features. | Ding Y; Wang J; Gao J; Fang Q; Li Y; Xu W; Wu J; Han D | 2021 | Jun | Sleep Breath | 25 | 2 | 787-795 |  | 10.1007/s11325-020-02168-0 |
| Single channel photoplethysmography-based obstructive sleep apnea detection and arrhythmia classification. | Chen X; Huang J; Luo F; Gao S; Xi M; Li J | 2022 |  | Technol Health Care | 30 | 2 | 399-411 |  | 10.3233/THC-213138 |
| Single-cell RNA-seq uncovers cellular heterogeneity and provides a signature for paediatric sleep apnoea. | Cortese R; Adams TS; Cataldo KH; Hummel J; Kaminski N; Kheirandish-Gozal L; Gozal D | 2023 | Feb | Eur Respir J | 61 | 2 |  |  | 10.1183/13993003.01465-2022 |
| Single-channel oximetry monitor versus in-lab polysomnography oximetry analysis: does it make a difference? | Behar JA; Palmius N; Zacharie S; Chocron A; Penzel T; Bittencourt L; Tufik S | 2020 | May | Physiol Meas | 41 | 4 | 44007 |  | 10.1088/1361-6579/ab8856 |
| Sleep Apnea Detection Using Multi-Error-Reduction Classification System with Multiple Bio-Signals. | Li X; Leung FHF; Su S; Ling SH | 2022 | Jul | Sensors (Basel) | 22 | 15 |  |  | 10.3390/s22155560 |
| Sleep Apnea Detection Using Wavelet Scattering Transformation and Random Forest Classifier. | Sharaf AI | 2023 | Feb | Entropy (Basel) | 25 | 3 |  |  | 10.3390/e25030399 |
| Sleep bruxism and its associations with insomnia and OSA in the general population of Sao Paulo. | Maluly M; Dal Fabbro C; Andersen ML; Herrero Babiloni A; Lavigne GJ; Tufik S | 2020 | Nov | Sleep Med | 75 |  | 141-148 | ClinicalTrials.gov/NCT00596713 | 10.1016/j.sleep.2020.06.016 |
| Sleep spindle activity in children with obstructive sleep apnea as a marker of neurocognitive performance: A pilot study. | Brockmann PE; Damiani F; Pincheira E; Daiber F; Ruiz S; Aboitiz F; Ferri R; Bruni O | 2018 | May | Eur J Paediatr Neurol | 22 | 3 | 434-439 |  | 10.1016/j.ejpn.2018.02.003 |
| Sleep staging from single-channel EEG with multi-scale feature and contextual information. | Chen K; Zhang C; Ma J; Wang G; Zhang J | 2019 | Dec | Sleep Breath | 23 | 4 | 1159-1167 | | 10.1007/s11325-019-01789-4 |
| SleepPPG-Net: A Deep Learning Algorithm for Robust Sleep Staging From Continuous Photoplethysmography. | Kotzen K; Charlton PH; Salabi S; Amar L; Landesberg A; Behar JA | 2022 | Nov | IEEE J Biomed Health Inform | PP |  |  |  | 10.1109/JBHI.2022.3225363 |
| Sleep-wake stage detection with single channel ECG and hybrid machine learning model in patients with obstructive sleep apnea. | Bozkurt F; UÃ§ar MK; Bilgin C; Zengin A | 2021 | Mar | Phys Eng Sci Med | 44 | 1 | 63-77 |  | 10.1007/s13246-020-00953-5 |
| Sleep-wake stages classification using heart rate signals from pulse oximetry. | Casal R; Di Persia LE; Schlotthauer G | 2019 | Oct | Heliyon | 5 | 10 | e02529 |  | 10.1016/j.heliyon.2019.e02529 |
| Snore-GANs: Improving Automatic Snore Sound Classification With Synthesized Data. | Zhang Z; Han J; Qian K; Janott C; Guo Y; Schuller B | 2020 | Jan | IEEE J Biomed Health Inform | 24 | 1 | 300-310 |  | 10.1109/JBHI.2019.2907286 |
| SomnNET: An SpO2 Based Deep Learning Network for Sleep Apnea Detection in Smartwatches. | John A; Nundy KK; Cardiff B; John D | 2021 | Nov | Annu Int Conf IEEE Eng Med Biol Soc | 2021 |  | 1961-1964 | | 10.1109/EMBC46164.2021.9631037 |
| Support vector machine prediction of obstructive sleep apnea in a large-scale Chinese clinical sample. | Huang WC; Lee PL; Liu YT; Chiang AA; Lai F | 2020 | Jul | Sleep | 43 | 7 |  |  | 10.1093/sleep/zsz295 |
| The Discovery, Validation, and Function of Hypoxia-Related Gene Biomarkers for Obstructive Sleep Apnea. | Wu X; Pan Z; Liu W; Zha S; Song Y; Zhang Q; Hu K | 2022 |  | Front Med (Lausanne) | 9 |  | 813459 |  | 10.3389/fmed.2022.813459 |
| The effect and relative importance of sleep disorders for all-cause mortality in middle-aged and older asthmatics. | Hu Z; Tian Y; Song X; Zeng F; Hu K; Yang A | 2022 | Nov | BMC Geriatr | 22 | 1 | 855 |  | 10.1186/s12877-022-03587-2 |
| The prediction of obstructive sleep apnea severity based on anthropometric and Mallampati indices. | Amra B; Pirpiran M; Soltaninejad F; Penzel T; Fietze I; Schoebel C | 2019 |  | J Res Med Sci | 24 |  | 66 |  | 10.4103/jrms.JRMS_653_18 |
| The Prediction of Obstructive Sleep Apnea Using Data Mining Approaches. | Manoochehri Z; Rezaei M; Salari N; Khazaie H; Khaledi Paveh B; Manoochehri S | 2018 | Oct | Arch Iran Med | 21 | 10 | 460-465 |  |  |
| The Predictive Role of Subcutaneous Adipose Tissue in the Pathogenesis of Obstructive Sleep Apnoea. | MolnÃ¡r V; Lakner Z; MolnÃ¡r A; TÃ¡rnoki DL; TÃ¡rnoki ÃD; Kunos L; TamÃ¡s L | 2022 | Sep | Life (Basel) | 12 | 10 |  |  | 10.3390/life12101504 |
| The Predictive Role of the Upper-Airway Adipose Tissue in the Pathogenesis of Obstructive Sleep Apnoea. | MolnÃ¡r V; Lakner Z; MolnÃ¡r A; TÃ¡rnoki DL; TÃ¡rnoki ÃD; Kunos L; Jokkel Z; TamÃ¡s L | 2022 | Oct | Life (Basel) | 12 | 10 |  |  | 10.3390/life12101543 |
| Towards Validating the Effectiveness of Obstructive Sleep Apnea Classification from Electronic Health Records Using Machine Learning. | Ramesh J; Keeran N; Sagahyroon A; Aloul F | 2021 | Oct | Healthcare (Basel) | 9 | 11 |  |  | 10.3390/healthcare9111450 |
| Transfer learning artificial intelligence for automated detection of atrial fibrillation in patients undergoing evaluation for suspected obstructive sleep apnoea: a feasibility study. | Gahungu N; Shariar A; Playford D; Judkins C; Gabbay E | 2021 | Sep | Sleep Med | 85 |  | 166-171 |  | 10.1016/j.sleep.2021.07.014 |
| Treatment usage patterns of oral appliances for obstructive sleep apnea over the first 60 days: a cluster analysis. | Sutherland K; Almeida FR; Kim T; Brown EC; Knapman F; Ngiam J; Yang J; Bilston LE; Cistulli PA | 2021 | Sep | J Clin Sleep Med | 17 | 9 | 1785-1792 | | 10.5664/jcsm.9288 |
| Upper airway effective compliance during wakefulness and sleep in obese adolescents studied via two-dimensional dynamic MRI and semiautomated image segmentation. | Choy KR; Sin S; Tong Y; Udupa JK; Luchtenburg DM; Wagshul ME; Arens R; Wootton DM | 2021 | Aug | J Appl Physiol (1985) | 131 | 2 | 532-543 |  | 10.1152/japplphysiol.00839.2020 |
| Usefulness of recurrence plots from airflow recordings to aid in paediatric sleep apnoea diagnosis. | Barroso-GarcÃ­a V; GutiÃ©rrez-Tobal GC; Kheirandish-Gozal L; Ãlvarez D; Vaquerizo-Villar F; NÃºÃ±ez P; Del Campo F; Gozal D; Hornero R | 2020 | Jan | Comput Methods Programs Biomed | 183 |  | 105083 |  | 10.1016/j.cmpb.2019.105083 |
| Using Topic Modeling to Develop Multi-level Descriptions of Naturalistic Driving Data from Drivers with and without Sleep Apnea. | McLaurin EJ; Lee JD; McDonald AD; Aksan N; Dawson J; Tippin J; Rizzo M | 2018 | Oct | Transp Res Part F Traffic Psychol Behav | 58 |  | 25-38 |  | 10.1016/j.trf.2018.05.019 |
| Using tracheal breathing sounds and anthropometric information for screening obstructive sleep apnoea during wakefulness. | Elwali A; Meza-Vargas S; Moussavi Z | 2019 | Feb | J Med Eng Technol | 43 | 2 | 111-123 |  | 10.1080/03091902.2019.1617799 |
| Utilisation of machine learning to predict surgical candidates for the treatment of childhood upper airway obstruction. | Liu X; Pamula Y; Immanuel S; Kennedy D; Martin J; Baumert M | 2022 | Jun | Sleep Breath | 26 | 2 | 649-661 |  | 10.1007/s11325-021-02425-w |
| Validation of the Withings Sleep Analyzer, an under-the-mattress device for the detection of moderate-severe sleep apnea syndrome. | Edouard P; Campo D; Bartet P; Yang RY; Bruyneel M; Roisman G; Escourrou P | 2021 | Jun | J Clin Sleep Med | 17 | 6 | 1217-1227 | ClinicalTrials.gov/NCT04234828 | 10.5664/jcsm.9168 |
| Vibration pattern recognition using a compressed histogram of oriented gradients for snoring source analysis. | Zhang Y; Zhao Z; Xu HJ; He C; Peng H; Gao Z; Xu ZY | 2020 |  | Biomed Mater Eng | 31 | 3 | 143-155 |  | 10.3233/BME-201086 |
| Vision Transformers (ViT) for Blanket-Penetrating Sleep Posture Recognition Using a Triple Ultra-Wideband (UWB) Radar System. | Lai DK; Yu ZH; Leung TY; Lim HJ; Tam AY; So BP; Mao YJ; Cheung DSK; Wong DW; Cheung JC | 2023 | Feb | Sensors (Basel) | 23 | 5 |  |  | 10.3390/s23052475 |
| Wavelet analysis of oximetry recordings to assist in the automated detection of moderate-to-severe pediatric sleep apnea-hypopnea syndrome. | Vaquerizo-Villar F; Ãlvarez D; Kheirandish-Gozal L; GutiÃ©rrez-Tobal GC; Barroso-GarcÃ­a V; Crespo A; Del Campo F; Gozal D; Hornero R | 2018 |  | PLoS One | 13 | 12 | e0208502 |  | 10.1371/journal.pone.0208502 |
| Wearable monitoring of sleep-disordered breathing: estimation of the apnea-hypopnea index using wrist-worn reflective photoplethysmography. | Papini GB; Fonseca P; van Gilst MM; Bergmans JWM; Vullings R; Overeem S | 2020 | Aug | Sci Rep | 10 | 1 | 13512 |  | 10.1038/s41598-020-69935-7 |
